# CT-based Rapid Triage of COVID-19 Patients: Risk Prediction and Progression Estimation of ICU Admission, Mechanical Ventilation, and Death of Hospitalized Patients

**DOI:** 10.1101/2020.11.04.20225797

**Authors:** Qinmei Xu, Xianghao Zhan, Zhen Zhou, Yiheng Li, Peiyi Xie, Shu Zhang, Xiuli Li, Yizhou Yu, Changsheng Zhou, Longjiang Zhang, Olivier Gevaert, Guangming Lu

## Abstract

The wave of COVID-19 continues to overwhelm the medical resources, especially the stressed intensive care unit (ICU) capacity and the shortage of mechanical ventilation (MV). Here we performed CT-based analysis combined with electronic health records and clinical laboratory results on Cohort 1 (n = 1662 from 17 hospitals) with prognostic estimation for the rapid stratification of PCR confirmed COVID-19 patients. These models, validated on Cohort 2 (n = 700) and Cohort 3 (n = 662) constructed from 9 external hospitals, achieved satisfying performance for predicting ICU, MV and death of COVID-19 patients (AUROC 0.916, 0.919 and 0.853), even on events happened two days later after admission (AUROC 0.919, 0.943 and 0.856). Both clinical and image features showed complementary roles in events prediction and provided accurate estimates to the time of progression (p<.001). Our findings are valuable for delivering timely treatment and optimizing the use of medical resources in the pandemic of COVID-19.

## Introduction

From 30 December through 11 October, the ongoing severe acute respiratory syndrome–coronavirus 2 (SARS-CoV-2) pandemic has caused over 37 million coronavirus disease 2019 (COVID-19) confirmed cases and 1 million deaths globally.^1^ The spread of COVID-19 continues to overwhelm the medical resources without effective therapeutics and vaccines. In particular, stressed intensive care unit (ICU) capacity and the shortage of mechanical ventilation (MV) are major factors that drive COVID-19 mortality rates.^2, 3, 4^ To enable sufficient supply of medical resources, rapid triage method for COVID-effected patients with potentially serious outcomes has become an urgent priority for reallocating medical resources as well as distributing patients to balance ICU loads across affected regions so as to deliver timely treatment.^5, 6, 7, 8^

Evaluating the severity of patients with infectious pneumonia have been applied in clinics such as measuring the acute physiology and chronic health evaluation II (APACHE-II) score and laboratory indicators including neutrophil-to-lymphocyte ratio (NLR).^9, 10, 11, 12^ However, the scoring systems of APACHE-II are highly subjective and time-consuming while laboratory indicators are not comprehensive enough to predict the adverse outcomes of the newly emerged COVID-19. Although computed tomography (CT) assessment by radiologists is now an important criterion for COVID-19 diagnosis and severity evaluation of COVID-19,^13^ it is limited by manual evaluation of radiologists with marked inter- and intra-observer variability and unable to provide accurate prognosis prediction. Better ways to utilize multi-modal data for grouping hospitalized COVID-19 patients according to their potential clinical outcomes remain to be developed to deliver specific treatment timely.

In this study, we provided risk stratification based on CT-based radiomics features and clinical data for COVID-19 patients in terms of stable or severe disease (requiring ICU) on admission. Then we developed specific outcome prediction (MV/ death) models for critically ill patients. Finally, we provided insights into estimating time to the progression (ICU/MV/death) for COVID-19 patients.

## Results

### Patient cohort

In total, 2362 patients were used in this study, including a primary cohort (Cohort 1, n = 1662) for model development included patients from 17 hospitals, and a validation cohort (Cohort 2, n = 700) consisted of patients from 9 external and independent medical centers (Figure 1, Figure S1, Table S1). Additionally, we built a specific subset of Cohort 2 (Cohort 3, n = 662) for patients from the 9 medical centers whose interval between admission and progression to critical outcomes (ICU/MV/death) were more than two days, aiming to evaluate the performance of our models on predicting events happening at least two days after admission. Prediction models were built for three prediction tasks, including ICU (adverse cases in Cohort 1/Cohort 2/Cohort 3, n = 96/59/21, respectively), MV (adverse cases in Cohort 1/2/3, n = 55/39/19), and death (adverse cases in Cohort 1/2/3, n = 31/28/20). Note that most patients with death were also in the MV group, while all patients with MV or death lay into the ICU group. In our study, 2207 patients (93.5%) were discharged without any adverse outcome (stable group), 155 (6.5%) patients developed adverse clinical outcomes and were admitted to the ICU (adverse group), of whom 94 (60.6%) required MV, and 59 (38.0%) died within 28 days from admission (Table 1, Table S2, Table S3). This cohort had 1229 men (52.0%) and 1133 women (48.0%), with a median age of 51.5 years (IQR, 39 - 64 years). The median age among men was 57 years (IQR, 45 - 68 years) and the median age among women was 52 years (IQR, 39 - 64 years). No statistical difference in age was found between men and women in this cohort.

**Table 1.** Clinical characteristics of COVID-19 patients in Cohort 1, Cohort 2, Cohort 3, and the whole cohort

|  | All patients (n = 2362) | Cohort 1 (n = 1662) | Cohort 2 (n = 700) | Cohort 3 (n = 662) | p-value |
| --- | --- | --- | --- | --- | --- |
| **Demographics** |  |  |  |  |  |
| Age (years) | 51.720 ± 15.646 | 52.465 ± 15.863 | 49.953 ± 14.984 | 48.985 ± 14.545 | < 0.001 |
| Gender (male) | 1229 (52.0%) | 881 (53.0%) | 348 (49.7%) | 338 (51.0%) | 0.143 |
| **Comorbidity** |  |  |  |  |  |
| Coronary heart disease | 172 (7.2%) | 123 (7.4%) | 49 (7.0%) | 36 (5.4%) | 0.732 |
| Chronic liver disease | 82 (3.4%) | 58 (3.4%) | 24 (3.4%) | 24 (3.6%) | 0.941 |
| Chronic kidney disease | 29 (1.2%) | 18 (1.0%) | 11 (1.5%) | 8 (1.2%) | 0.325 |
| COPD | 51 (2.1%) | 33 (1.9%) | 18 (2.5%) | 12 (1.8%) | 0.371 |
| Diabetes | 261 (11.0%) | 191 (11.4%) | 70 (10.0%) | 59 (8.9%) | 0.291 |
| Hypertension | 500 (21.1%) | 370 (22.2%) | 130 (18.5%) | 110 (16.6%) | 0.045 |
| Carcinoma | 61 (2.5%) | 44 (2.6%) | 17 (2.4%) | 15 (2.2%) | 0.759 |
| **Clinical symptom** |  |  |  |  |  |
| Fever | 1950 (82.5%) | 1340 (80.6%) | 610 (87.1%) | 580 (87.6%) | < 0.001 |
| Cough | 1651 (69.8%) | 1170 (70.3%) | 481 (68.7%) | 455 (68.7%) | 0.416 |
| Myalgia | 553 (23.4%) | 467 (28.0%) | 86 (12.2%) | 78 (11.7%) | < 0.001 |
| Fatigue | 952 (40.3%) | 719 (43.2%) | 233 (33.2%) | 224 (33.8%) | < 0.001 |
| Headache | 191 (8.0%) | 138 (8.3%) | 53 (7.5%) | 50 (7.5%) | 0.551 |
| Nausea or vomiting | 116 (4.9%) | 84 (5.0%) | 32 (4.5%) | 30 (4.5%) | 0.620 |
| Diarrhea | 167 (7.0%) | 115 (6.9%) | 52 (7.4%) | 48 (7.2%) | 0.659 |
| Abdominal pain | 28 (1.1%) | 21 (1.2%) | 7 (1.0%) | 6 (0.9%) | 0.589 |
| Dyspnea | 403 (17.0%) | 312 (18.7%) | 91 (13.0%) | 70 (10.5%) | 0.001 |
| **Outcome** |  |  |  |  |  |
| ICU | 155 (6.5%) | 96 (5.7%) | 59 (8.4%) | 21 (3.1%) | 0.017 |
| MV | 96 (3.9%) | 55 (3.3%) | 39 (5.5%) | 19 (2.8%) | 0.010 |
| Death | 59 (2.4%) | 31 (1.8%) | 28 (4.0%) | 20 (3.0%) | 0.002 |
| **\* The mean interval (d) (IQR)** |  |  |  |  |  |
| Admission - ICU | 4.4 (1 - 6) | 4.6 (1 - 6) | 4.2 (1 - 6.5) | 8.4 (5 - 10.5) | 0.207 |
| Admission - MV | 6.1 (2 - 9) | 6.1 (1 - 10) | 6.1 (2 - 8.25) | 9.6 (5 - 13.5) | 0.758 |
| Admission - death | 16.1 (9.5 - 21.5) | 16.5 (9.5 - 24) | 15.6 (10 - 18) | 15.9 (11.8 - 18.3) | 0.386 |
| Admission - discharge | 15.7 (7 - 22) | 13.3 (5 - 9) | 19.3 (13 - 25) | 19.3 (13 - 25) | < 0.001 |

Note. P-values show statistically significant differences in features between Cohort 1 and Cohort 2. There were statistically significant differences in prognostic features (e.g. age, dyspnea) in Cohort 1 and Cohort 2, but there was no significant difference in these features of positive cases (refers to the adverse group where patients required ICU admission) in the two cohorts (**Table S3**). Thus, this difference may be due to the discrepancy in the proportion of Hubei cases (Cohort 1, 69.8%; Cohort 2, 80.1%), which have a higher proportion of severe outcomes (6.9%, 8.6%, respectively).

**Figure 1.**
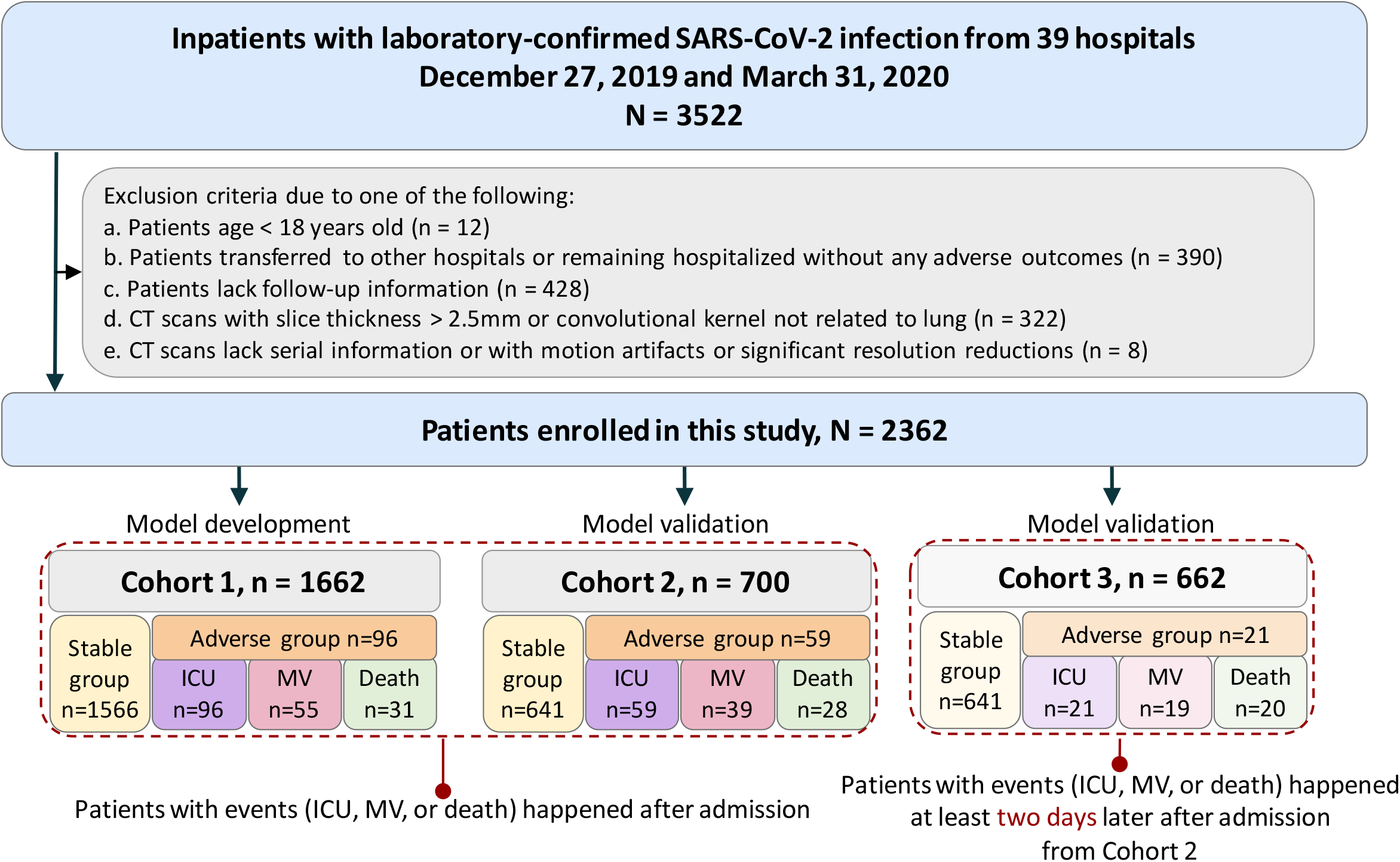
Illustration of workflow in this study. **(a)** Our primary cohort (Cohort 1, n = 1662) for model development included patients from 17 hospitals, and our validation cohort (Cohort 2, n = 700) consisted of patients from 7 external and independent medical centers. Additionally, we built a specific cohort (Cohort 3, n = 662) for patients from the 7 medical centers whose interval between admission and progression to critical outcomes (ICU/MV/death) were more than two days, aiming to evaluate the performance of our models on predicting events happening at least two days after admission. **(b) Explanation of our data split and the corresponding usages**. (1) Step one: feature visualization of Cohort 1 and Cohort 2 to get the preliminary intuitive sense; (2) Step two: 70% samples of Cohort 1 were picked as the training set using stratified sampling based on death cases, where 5-fold cross-validation was used to tune the hyperparameters of the models; (3) Step three: model selection was performed on the remaining 30% samples of Cohort 1; (4) Step four: Cohort 2 and Cohort 3 were used to evaluate model performance in different aspects.

### Comparison of radiomics models with other modalities

We recognized the marked differences of CT-based radiomics data (abbreviated as Radiom), Clinical records (abbreviated as Clin), Laboratory results (abbreviated as Lab), and Radiologists’ semantic data (abbreviated as R-score) on Cohort 1 and Cohort 2 between negative outcome patients and positive outcome patients (Figure 2, Figure S3, Table S2, Supplementary Appendix 1). The optimal models for each data type (i.e. Radiom, RadioClin, RadioClinLab, ClinLab, and R-score) were chosen on Cohort 1 and validated on Cohort 2 and Cohort 3 (Table 2, S4-5, Figure 3). On Cohort 2, radiomics features alone (Radiom) showed good performance to predict ICU (AUROC 0.869, AUPRC 0.441), MV (AUROC 0.805, AUPRC 0.245), and death (AUROC 0.667, AUPRC 0.136). When combined with clinical features (RadioClin), the performance of models improved significantly (all three events p-value < .001) (Table S4-5). Notably, as we continued to add the lab results (RadioClinLab), models achieved optimal performance on all three events (AUROC ICU: 0.916, MV: 0.919, death: 0.853; AUPRC ICU: 0.563, MV: 0.476, death: 0.248). RadioClinLab models also outperformed clinical data alone models (ClinLab) (all three events p-value < .001) (Table S4-5, Figure S4), suggesting the importance of radiomics features in predicting severe outcomes. Similarly, RadioClinLab models also had comparable performance on Cohort 3 for ICU (AUROC 0.919, AUPRC 0.348), MV (AUROC 0.943, AUPRC 0.388), and death (AUROC 0.856, AUPRC 0.218). These results demonstrated the models’ ability to predict severe events that occur at least two days after admission (Table 2, Table S4).

**Table 2.**
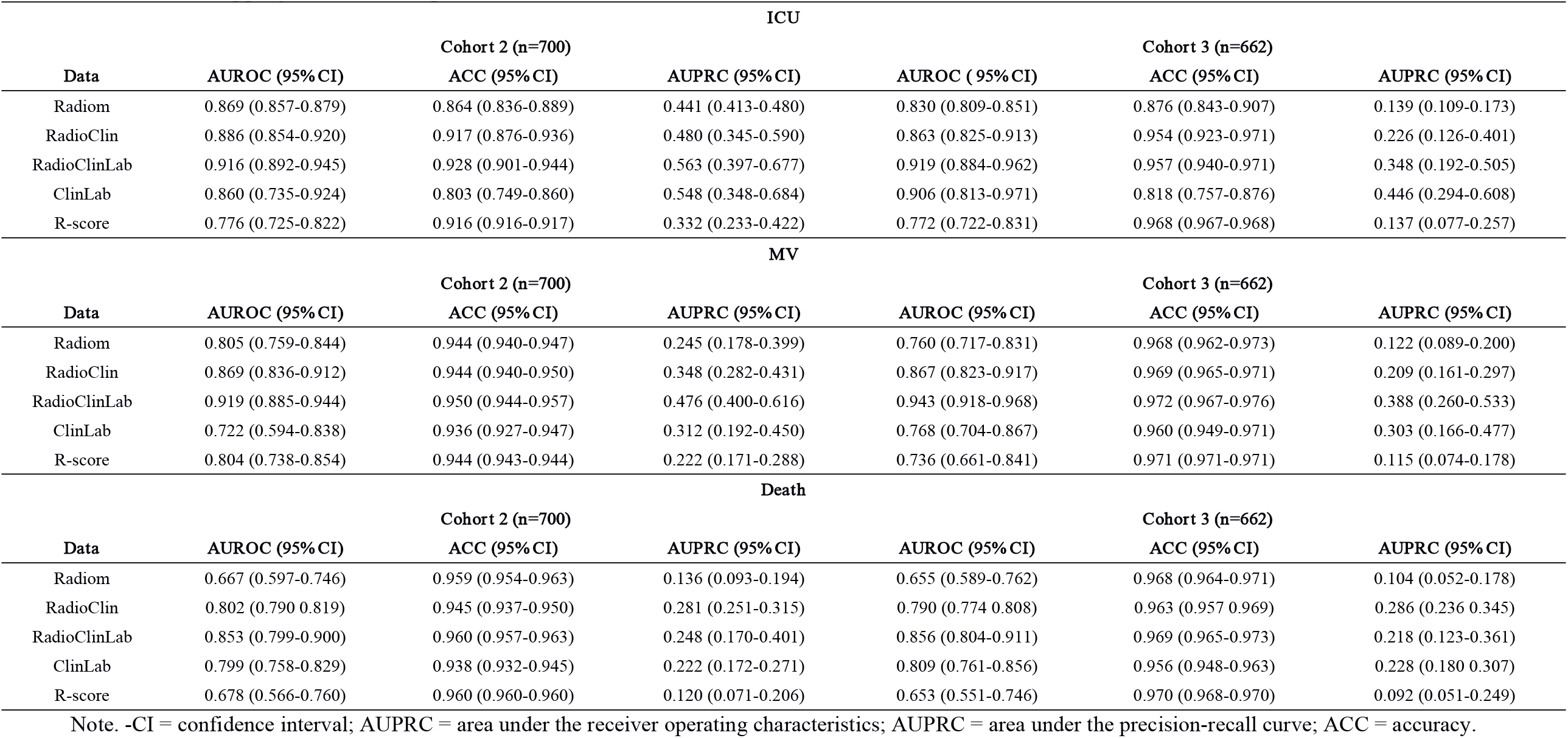
Bootstrapping results of the optimal models in Cohort 2 and Cohort 3

**Figure 2.**
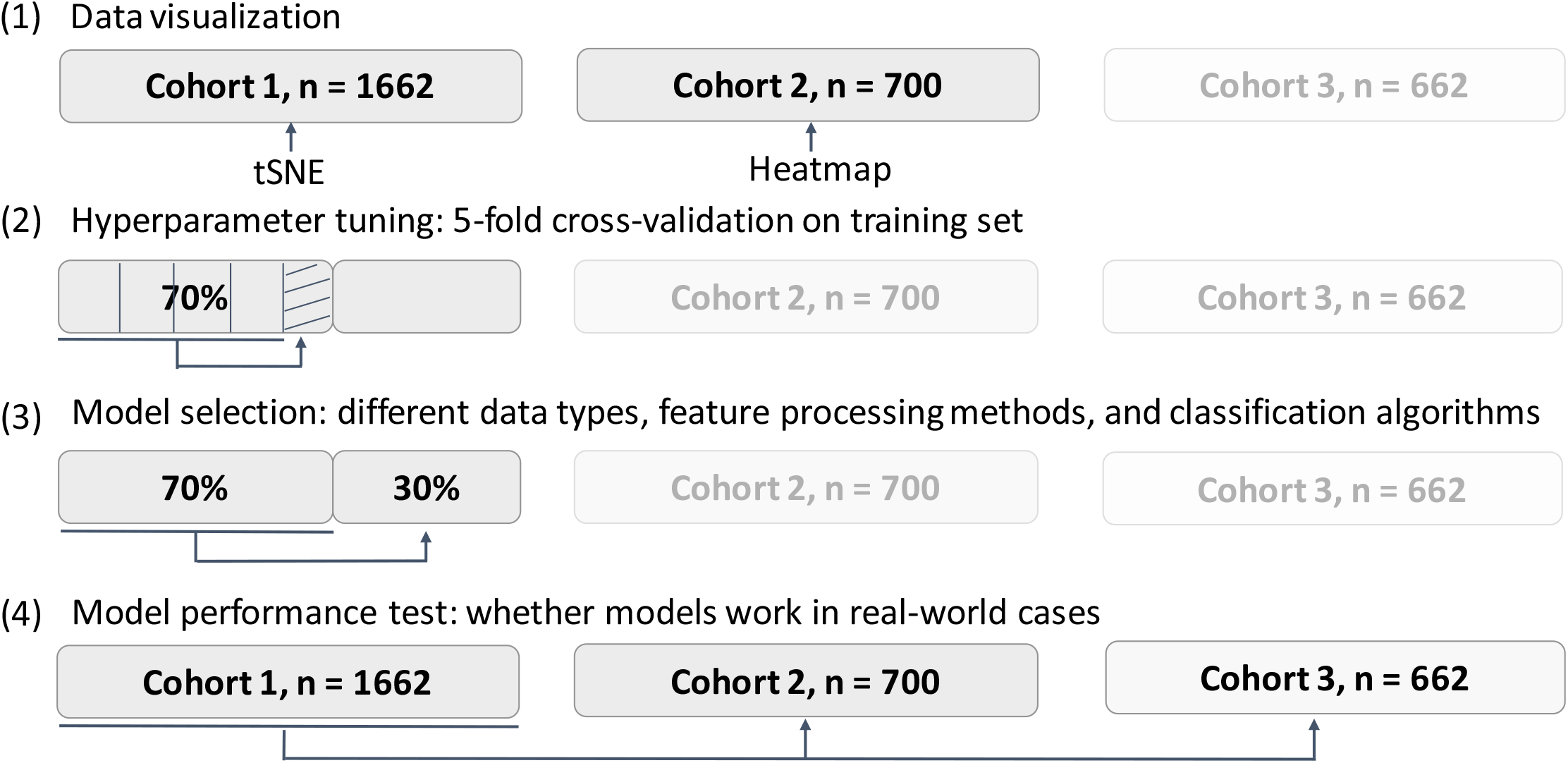
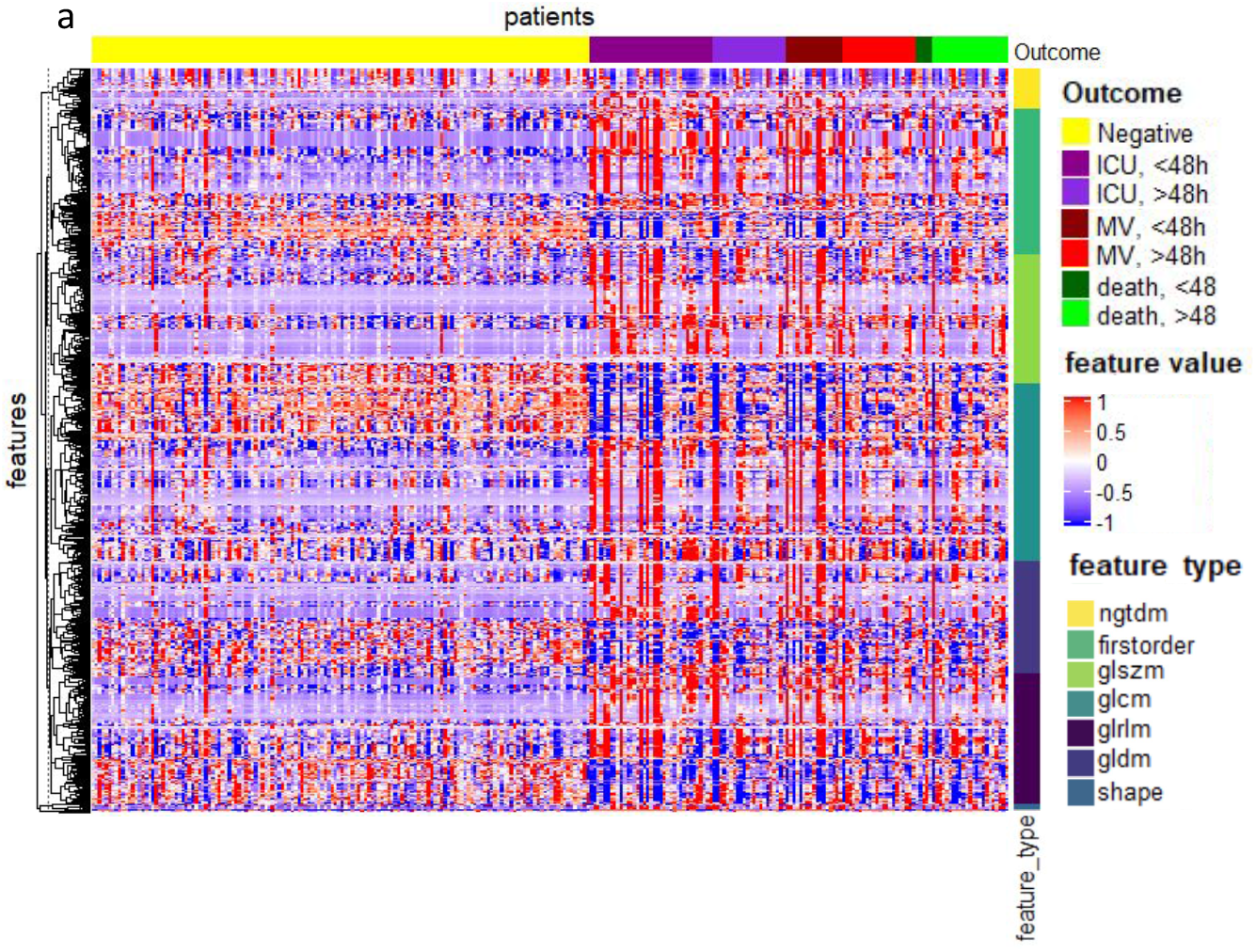
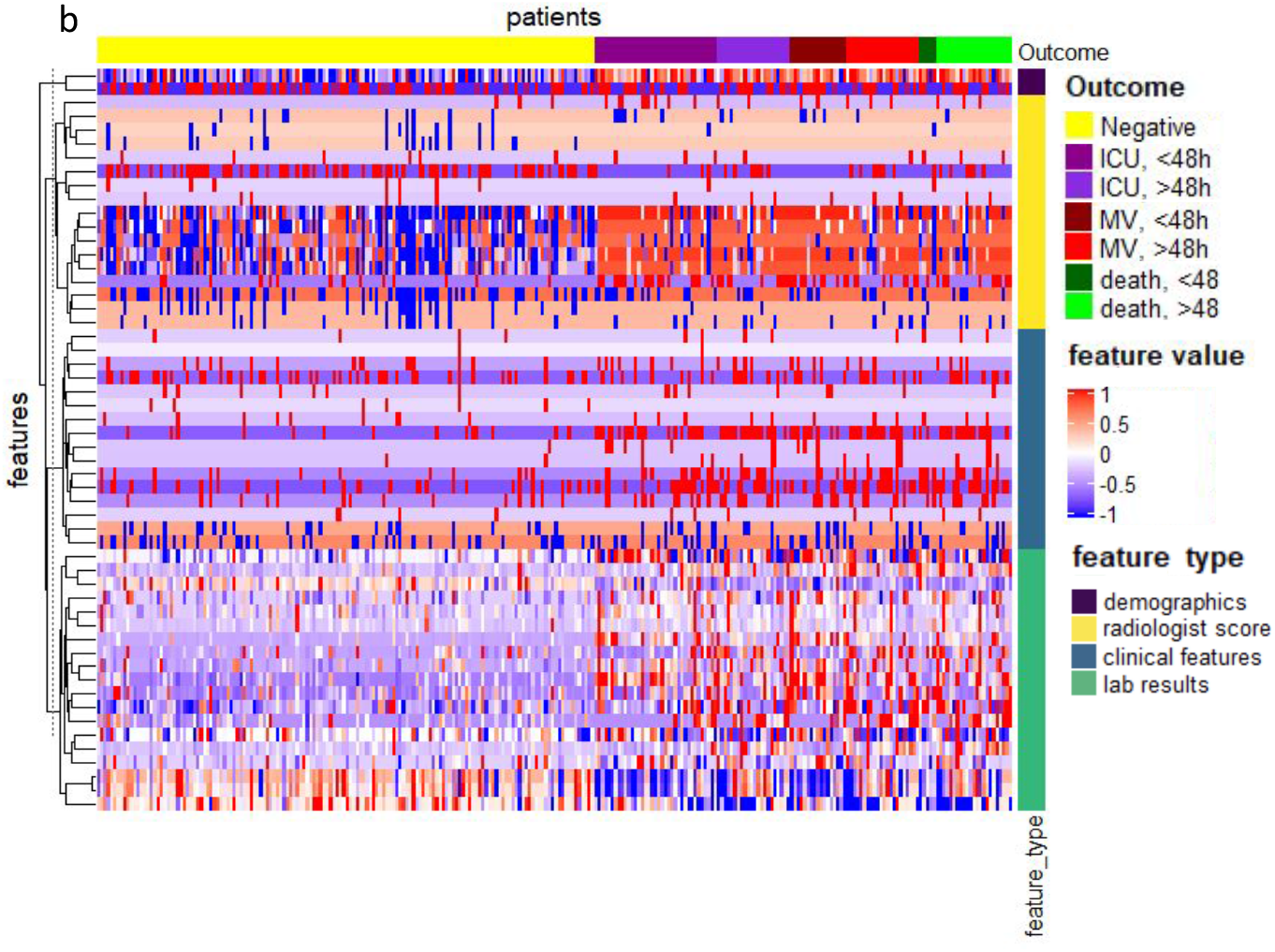
Heatmap showing the prognostic performance of (a) radiomics data and (b) clinical data and R-score data on Cohort 2 with clustering of features. 150 Negative patients were randomly selected as well as all patients having outcomes of ICU admission, Mechanical Ventilation or Death to draw the heatmap. For patients with more than one adverse outcome, they will appear as samples in each corresponding category. The patients were grouped based on adverse outcomes (i.e. ICU admission, MV, and death) and whether the event occurred within 48 hours after admission. The features were clustered within their categories to better visualize the data. The differences between negative outcome patients (yellow) and positive outcome patients can be seen from both (a) and (b), with some features showing different patterns for negative (patients discharged without any adverse outcomes) or positive patients (patients who required ICU, MV, or death while hospitalized). Almost all CT image features showed good discrimination between negative and severe outcome patients and had more obvious distinctions compared to clinical data. Among clinical data, lab results and demographics had good discriminating power. Part of radiologists’ score features had good discriminating power while clinical features have comparatively weak discriminating power. Regarding the distinctions between ICU admission, mechanical ventilation, and death, CT image features showed better discriminating power than clinical data. In CT image features, from ICU to MV to death, trends of value increasing or decreasing can be observed while in clinical data, this kind of trend is not visible.

**Figure 3.**
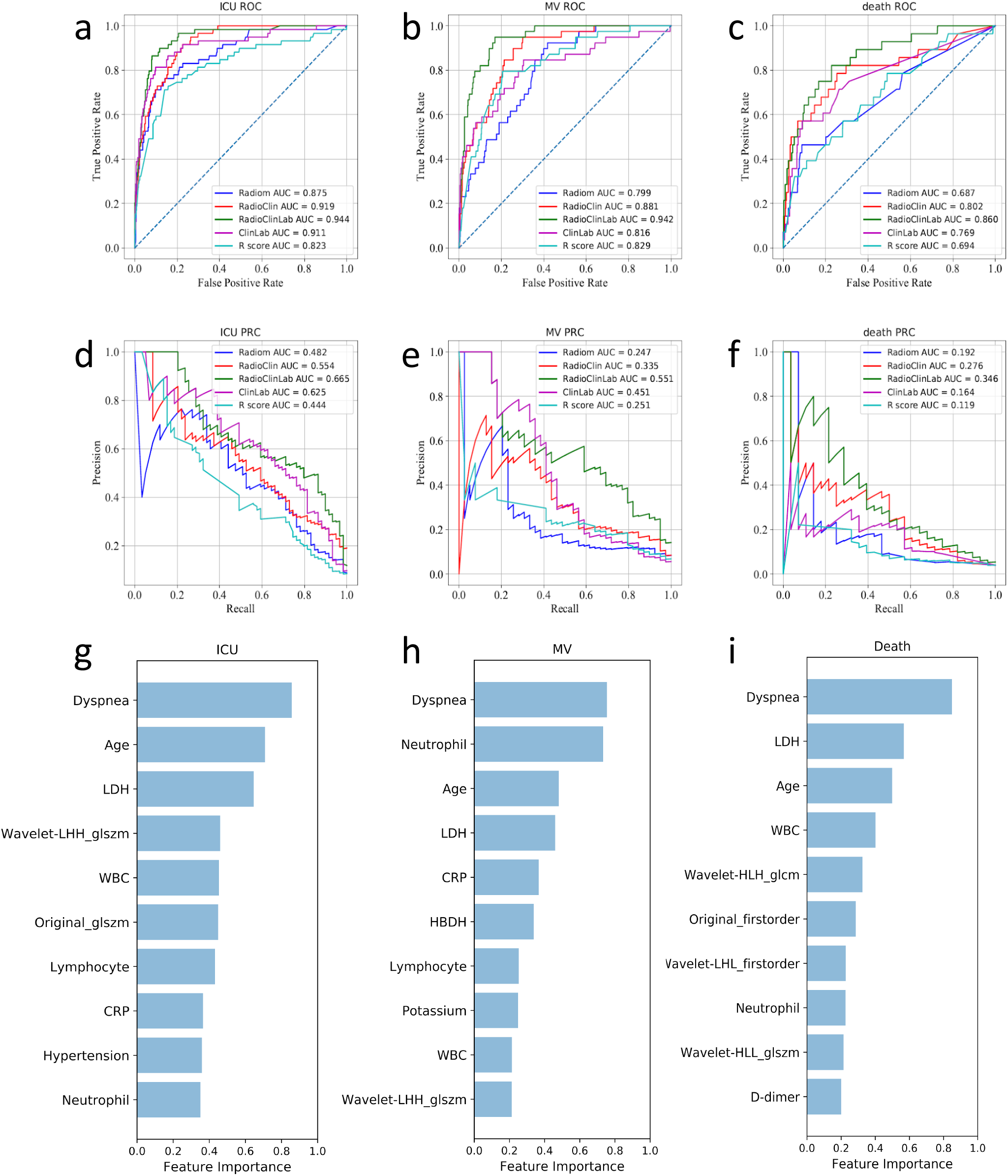
The model performances in the prediction of three outcomes (Cohort 2) and the ten most important features in the three outcome prediction tasks. The first and second row presented ROC curves and PR curves for predicting three events of models based on different data types. a) and d), b) and e), c) and f) indicated that RadioClinLab based models for predicting ICU/MV/death achieved the highest AUROC (0.944/0.942/0.860) and AUPRC (0.665/0.551/0.346), respectively. g-i) The ten most important features and their relative importance based on thirty bootstrapping experiments for the three prediction tasks based on the feature importance of the LightGBM classifiers.

### Comparison of radiomics with radiologists’ scoring

The performance of Radiom models was overall superior to that of radiologist score (R-score) models on both two validation cohorts for the three tasks (ICU/MV/death: Cohort 2 AUROC 0.776/0.804/0.678, AUPRC 0.332/0.222/0.120; Cohort 3 AUROC 0.772/0.736/0.653, AUPRC 0.137/0.115/0.092) (Table 2). Specifically, Radiom models had significantly improved predictive value in predicting ICU (p < .001) and were comparable to R-score models with a higher AUPRC for MV (p = .003) and death (p = .021) on Cohort 2. The predictive value of Radiom for ICU and MV happening two days later was higher than R-score, while there was no significant difference between these two models on prediction of death on Cohort 3 (Table S4-5, Figure S4).

### Key imaging features and clinical prognostic indicators

Among the top-ranking prognostic indicators, clinical data and radiomics features showed a complementary role with no significant correlations (Figure 3, Figure S5-6). In clinical data, elder age, dyspnea, higher lactate dehydrogenase (LDH) and inflammatory factors (white blood cell (WBC), neutrophil) signaled severe outcomes. Particularly, hypertension and some inflammatory factors (lower lymphocyte, higher C-reactive protein (CRP) and neutrophil)) were valuable for predicting ICU admission, also higher potassium and α-Hydroxybutyrate dehydrogenase (HBDH) and several inflammatory factors (lower lymphocyte, higher CRP) were predictive for MV, while higher D-dimer provided great diagnostic value for death. Most clinical variables were independently correlated with disease progression (Supplementary Appendix 5). Furthermore, GLSZM-based, GLCM-based, and first-order radiomics features are important features for the prediction of outcomes. In addition, our R-score model suggested that diffuse pulmonary parenchymal ground-glass and consolidative pulmonary opacities in the left upper lobe and pleural effusion increased the adverse outcomes (ICU, MV, death) in COVID-19 patients. Notably, crazy-paving on the initial CT chest was a risk factor of death. (Table S6, Figure S7)

### Individual severe-event-free survival analysis and performance of time-to-event models

Next, we used time-to-event modeling to stratify survival outcomes of patients. We first separated the patients into high-risk and low-risk groups and evaluated the survival curves of the two groups. Kaplan-Meier curves using the predicted score with the optimal RadioClinLab were generated (Figure 4). The high-risk group (ICU: 40 observations with 18 events, MV: 23 observations with 8 events, death: 13 observations with 3 events) had a much lower survival probability compared to the low-risk group (ICU: 642 observations with 32 events, MV: 659 observations with 28 events, death: 669 observations with 19 events) in all 3 tasks with a significant statistical difference (p < 0.001, log-rank test).

**Figure 4.**
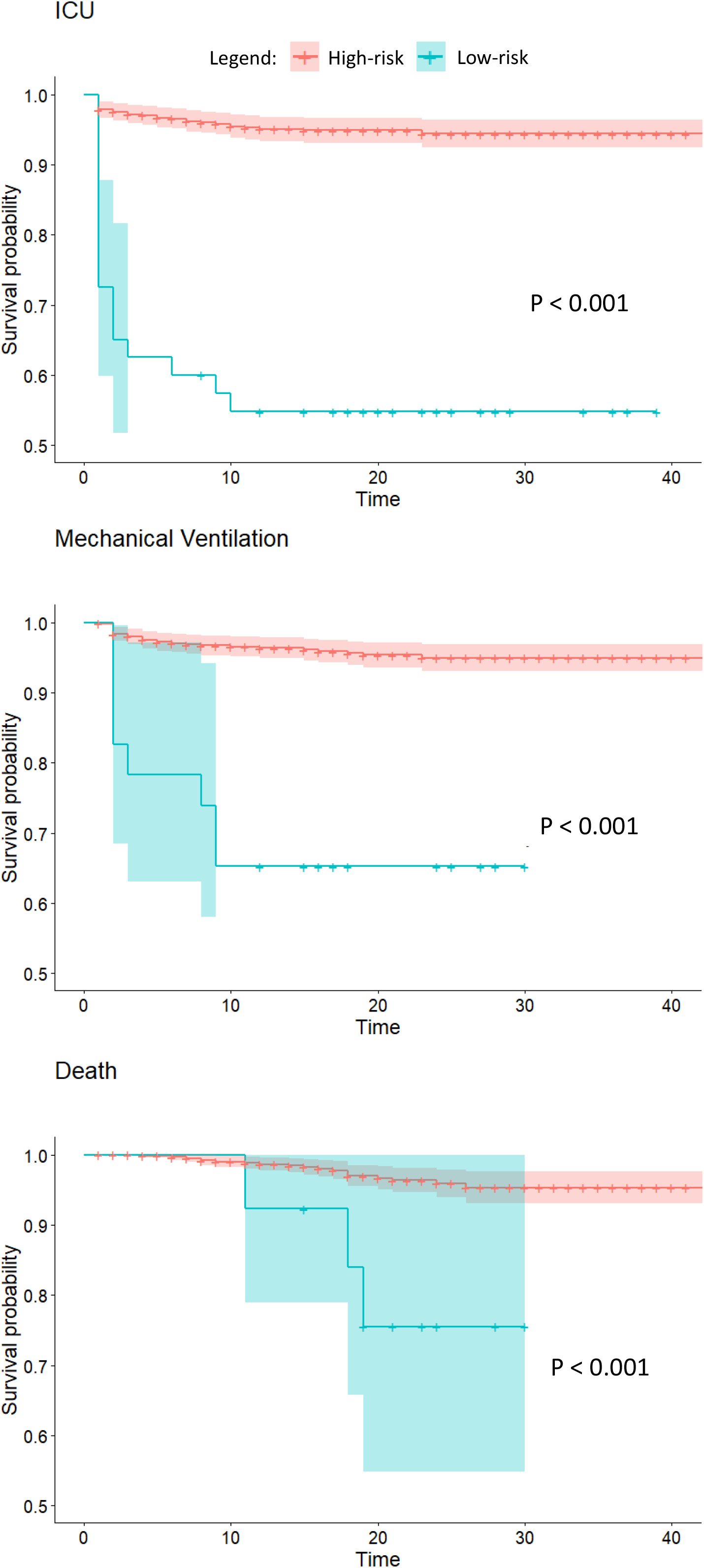
Kaplan-Meier curves for 3 tasks in Cohort 2. Risk groups were divided according to model predicted scores. (a) ICU admission (b) mechanical ventilation, and (c) death (high-risk: risk=1, low-risk: risk=0)

According to the results of time-to-event prediction (Table S9) on Cohort 2, the RadioClinLab showed the highest concordance index values on three prediction tasks (0.917, 0.888, and 0.906). Additionally, the RadioClinLab outperformed other models on ICU and MV prediction (Brier score 0.061 and 0.053) while the ClinLab model performed best on death prediction (Brier score 0.028). On Cohort 3, RadioClinLab showed the highest concordance index values on three tasks: 0.921, 0.884 and 0.911 and the lowest integrated Brier score on ICU and MV prediction: 0.039 and 0.036 while the ClinLab model showed the lowest integrated Brier score of 0.027. The bootstrapping experiments (Table S10) showed that on Cohort 2, RadioClinLab showed the highest concordance index on three tasks (p<0.001, paired one-sided t-test) and the lowest integrated Brier score on ICU and MV prediction (p<0.03) while there was no statistically significant difference in the integrated Brier score values between RadioClinLab and ClinLab on death prediction. Generally, these results showed that Radiom, RadioClinLab and ClinLab models achieved satisfactory performances in time-to-event prediction. In particular, the combination of radiomics features and clinical data contributed most to the prediction and provided the most accurate estimates to the time in days that critical care demands are required.

## Discussion

Our study achieved three goals. First, we provided risk stratification based on CT-based radiomics features and clinical data for COVID-19-infected patients in terms of stable or severe disease (requiring ICU) on admission. Second, our models provided specific outcome prediction (MV and death) for critically ill patients. Finally, we offered insights into estimating time to progression of severe events (i.e. ICU, MV, and death). This analysis potentially enables rapid stratification and timely intensive care management of patients during this pandemic.

We carefully defined outcome events (i.e. ICU, MV, or death) as prediction labels rather than the general risk severity, so that different medical centers can optimize the resources allocation by utilizing the prediction outcomes. According to our prognosis estimation results, it is possible to request medical resource transfers, such as personnel, local ICU beds, or MV from the Emergency Medical Services command as well as distribution of stable patients from overloaded local ICUs to neighboring affected regions with lower COVID-19 prevalence to balances ICU loads. Additionally, the prediction of MV on admission allows for closer monitoring and repeat assessments of patients over time to determine priority for initiating MV, because there is typically only a limited time window for life saving when their breathing deteriorates.^20^ Furthermore, combining predictions of demand for medical resources with outcome estimation of death anticipated the need to allocate resources to the patients who are most likely to benefit, which may also help develop priority rationing strategies during pandemics.^21^

Our findings demonstrated the predictive value of CT-based imaging for outcome predictions of CVOID-19 patients. The performance of radiomics-based models (Radiom) was better than radiologist’s scores (defined as R-score). Concretely, we found that first-order texture, and higher-order radiomics features (i.e. GLSMZ and GLCM-based) constituted the most important predictors. Our results also indicated that the feature values of diffuse pulmonary parenchymal ground-glass and consolidative pulmonary opacities in the left upper lobe as well as pleural effusion increased the adverse outcomes (ICU, MV, death) in COVID-19 patients, which were consistent with prior findings.^22, 23, 24^ Additionally, crazy paving was a predictor of death.^25^

Among the identified clinical predictors in our study, elder age, dyspnea, a liver biochemistry marker (higher lactate dehydrogenase (LDH)) were significant in all three prediction tasks.^26, 27, 28, 29^ Furthermore, the change of various inflammatory factors (higher white blood cell (WBC), C-reactive protein (CRP) and neutrophil, and lower lymphocytes) was predictive for the three severe events, consistent with current research that SARS-CoV-2 may accelerate the inflammatory response and cause the fluctuation of inflammatory factors, thereby leading to severe immune injury and lymphopenia.^26, 27, 30, 31, 32, 33^ Previous studies also indicated that leukocytosis resulting from a mixed infection of bacteria and fungi in the context of viral pneumonia indicates poor outcomes.^34, 35^ In addition, our study suggests that electrolyte and acid-base balance (K+) relating to respiratory function and the indicator of myocardial infarction (higher α-Hydroxybutyrate dehydrogenase (HBDH)) contributes to the prediction of progression to severe illness requiring MV, while D-dimer was associated with an increased risk of in-hospital mortality, in agreement with previous studies.^11, 12, 26, 32, 36^ Other features such as comorbidity (e.g. hypertension) were also related to poor prognosis.^27, 29^

Our work has several limitations. First, we did not consider the effect of different treatments on the prognosis of patients among clinical centers. In our study, several treatments were adopted including oxygen therapy, MV, ECMO, antiviral treatment, antibiotic treatment, glucocorticoids, and intravenous immunoglobulin therapy. In-depth comparison of different treatment outcomes might improve response prediction. Second, ten well-experienced thoracic radiologists analyzed the CT images in consensus and evaluated traditional imaging features in our study, however, we did not study inter-reader variability and such an analysis might need to be addressed in future work. In addition, although our study had a large sample size with clear prognosis information, the numbers of endpoints were limited and only from Chinese hospitals which could potentially limit the generalizability of models in other areas. Finally, additional validation across populations from European and American hospitals are needed to further validate the reported models.

In conclusion, we developed computational models with clinical prognostic estimation functions incorporating CT-based radiomics features as well as clinical data from electronic medical records for COVID-19 patients. This information may aid in delivering proper treatment and optimizing the use of limited medical resources in the current pandemic of COVID-19.

## Methods

### Patient cohort

Our data in this study was collected from 39 hospitals in China. All patients (n = 3522) followed the inclusion criteria: (a) confirmed positive SARS-CoV-2 nucleic acid test; (b) chest CT examinations and laboratory tests on the date of admission; (c) clear short-term prognosis information was available (discharge, or adverse outcomes including the admission to ICU, requiring MV support, and in-hospital death). After screening with exclusion criteria, 2363 from 26 medical centers were analyzed in our study (Figure 1, Figure S1, Table S1). This study protocol was approved by the institutional review board of Jinling Hospital, Nanjing University School of Medicine (2020NZKY-005-02).

### Data collection and processing

Our multi-modal data for each patient included (a) Clinical records (abbreviated as Clin): demographics, comorbidities, and clinical symptoms; (b) Laboratory results (abbreviated as Lab): blood routine, blood biochemistry, coagulation function, infection-related biomarkers; (c) CT-based radiomics features (abbreviated as Radiom); (d) Radiologists’ semantic data (abbreviated as R-score); (e) Time-to-event data: the time intervals between the date of admission and the date of development of adverse outcomes (requiring ICU, MV, and death), or the date of discharge. (Table S2, Supplementary Appendix 1) To address the imbalance and high feature dimensionality in modeling, we adopted several combinations of methods to downsample the negative cases (n=2207, referred to the stable group where patients discharged without any adverse outcome) and oversampling the positive cases (n=155, referred to the adverse group where patients required ICU admission, including 94 patients who needed MV and 59 death) to enhance models’ learning ability for the imbalanced data (Supplementary Appendix 2).

### Model development and prediction evaluation

There were three binary classification tasks in this study, namely, stable (negative) samples vs. adverse (ICU) samples, non-MV samples vs. MV samples, and survival samples vs. death samples. To test the prediction performances of different data type combinations, multivariable models based on five types of data were developed and compared: 1) radiomics data only (denoted as “Radiom”); 2) radiomics, clinical features (including demographics, comorbidity and clinical symptoms) (denoted as “RadioClin”); 3) radiomics data, clinical features, and laboratory results data (denoted as “RadioClinLab”); 4) clinical features and laboratory results (denoted as “ClinLab”); 5) radiological score based on the linear combination of semantic imaging features evaluated by radiologists (denoted as “R-score”). To confirm the patients were reasonably grouped based on the adverse outcomes and whether the event occurred within 48 hours, we first provided an intuitive manner to understand the distribution of all types of features used in this study with the help of heatmaps and t-distributed Stochastic Neighbor Embedding (t-SNE) in terms of ICU, MV, and death (Supplementary Appendix 3).

To systematically explore the performance of multiple machine-learning classifiers, we used the following approaches to predict outcomes: 1) Logistic Regression (LR); 2) Random Forest (RF); 3) Support Vector Machine (SVM); 4) Multilayer Perceptron (MLP); 5) LightGBM. In Cohort 1 (n=1662), the data were split into training and testing sets (ratio 7:3) using a stratified random sampling based on death cases. We used 5-fold cross-validation on the training set only to tune the model hyperparameters (Supplementary Appendix 4). Both a randomized search with accuracy as the optimization goal and a grid search with F1 score as the optimization goal were implemented on the 5-fold cross-validation and the predictive performances were evaluated on the test set of Cohort 1. Finally, to select an optimal model for each prediction task, five models with the top receiver operating characteristic (AUROC)^14^ were firstly selected and the model with the highest precision-recall (AUPRC)^15^ curves was then chosen as the optimal model for each outcome prediction because AUROC and AUPRC could show model accuracy, precision and recall in a more comprehensive manner with varying thresholds.

### Model validation and comparison

We tested the statistical difference of the performance of selected models with 30 bootstrapped resamples on unseen data (Cohort 2 n = 700, Cohort 3 n = 662, Figure 1) and used the AUROC and AUPRC curves to estimate their generalization ability. Particularly, with Cohort 3, we could verify models’ ability of predicting events that will occur two days later, which may allow the healthcare system to have at least two days to plan ahead and react to the demand for resources. Box plots were also drawn to compare the performances of the optimal models found based on Cohort 1 in three classification tasks. Finally, we selected an optimal model for each prediction task based on the results of the paired one-sided t-test, which compared the AUROC and AUPRC of models consisting of different data types (Radiom, RadioClin, RadioClinLab, ClinLab). Additionally, we constructed the R-score model using logistic regression based on semantic features to compare with the Radiom model (on both Cohort 2 and Cohort 3) and found out the traditional image features that were helpful to predict the outcome events (Supplementary Appendix 2).

### Analysis of predictive features

We identified the feature importance from the selected optimal models and normalized the highest importance scores in each of the bootstrapping experiments on Cohort 2 (n =700). By taking an average of the feature importance values over thirty bootstrapping experiments, we then focused on the ten most important features for each prediction task. We also plotted the pairplot of the most important features to visualize the relationship of top ten features. Furthermore, we performed the independent two-sided t-test (continuous variables, with normal distribution), proportional z-test (categorical variables) and rank sum test (continuous variables, without normal distribution) to validate the statistical significance in the feature values of positive cases and all cases in Cohort 1, Cohort 2 after firstly using Shapiro Wilk normality test.

### Time-to-event modeling

Cox regression with the l1 penalty and scikit-survival package 0.12.1 was adopted on time-to-event data in Cohort 1 (n = 1277, 77% of the patients originally in Cohort 1 had event time recorded) and Cohort 2 (n = 682, 97% of the patients originally in Cohort 2 had event time recorded).^16, 17, 18, 19^ Three different data combinations were used for the time-to-event modeling: Radiom, RadioClinLab, and ClinLab. We used five-fold cross-validation on Cohort 1 to determine the “alpha_min_ratio” hyperparameter,^18, 19^ and calculated the performance on Cohort 2. We used the concordance index (C index) and the integrated Brier score to evaluate the models. On Cohort 1, the optimal model for each data combination was chosen in a similar manner as previously described for the classification tasks by first filtering based on mean C index and then optimizing the mean integrated Brier score on the three tasks. Next, we used Kaplan-Meier analysis to visualize the time-to-event models and the log-rank test to estimate significance. A “high-risk” and “low-risk” group was created according to the predicted score for each patient on each task with the optimal RadioClinLab model. To group the patients into the high-risk group and the low-risk group, we first calculated the ratios of positive cases in Cohort 1, then set thresholds on the predicted probability of the test samples to separate patients according to the ratios based on Cohort 2.

### Statistical analysis

SPSS v15.0 [Chicago, SPSS Inc.] and MedCalc statistical software were used for statistical analysis. The Shapiro-Wilk test was used to evaluate the normality of quantitative data among the selected top important features. Mean and standard deviation (SD) were used to describe normally distributed data, while the median and interquartile range (IQR) was used to describe non-normally distributed data. Categorical variables were presented as numbers and percentages. The AUROC, AUPRC, accuracy value and their 95% CI were listed to assess the model performance. The paired one-sided t-test was used to calculate the statistical significance of the difference between each AUROC and AUPRC value in the bootstrapping experiments. Chi-square test and Fisher’s exact test were exploited to compare categorical data while independent t-test and Wilcoxon rank sum test were used to compare the feature values of continuous variables in positive and negative cases in the entire cohort (n = 2362). Proportional test was done to compare the feature values of categorical variables in positive and negative cases among the most important features found by classifiers and test the statistical significance of categorical variables between Cohort 1 and Cohort 2. Kaplan-Meier survival analysis was done on the high-risk and low-risk group based on predictions and log-rank test was used to evaluate statistical significance.

## Data Availability

The data that support the findings of this study are available on request from the corresponding author (G.M.L.). The data with participant privacy/consent are not publicly available due to hospital regulation restrictions.

## Acknowledgements

This study could not have been possible without expertise from a variety of teams for data collection and model development. We would like to acknowledge suggestions from Dr. Song Luo, Dr. Zhao Shi, Dr. Pritam Mukherjee, Dr. Heather Marie Selby, and Dr Mu Zhou. This work was supported by National Basic Research Program (973 Program) (grants No. 2014CB744504 to Guangming Lu) and the National Institute of Biomedical Imaging and Bioengineering of the National Institutes of Health (NIBIB https://www.nibib.nih.gov/), R01 EB020527 and R56 EB020527 to OG.

## Author information

### Author notes

These two authors contributed equally: Qinmei Xu, Xianghao Zhan; Corresponding authors: Guangming Lu and Olivier Geveart.

## Contributions

Q.M.X., X.H.Z., G.M.L., and O.G. conceived the study. Q.M.X.and X.H.Z. reviewed the literature. Q.M.X., P.Y.X., C.S.Z., L.J.Z., and G.M.L. provided clinical expertise. Q.M.X. and C.S.Z. collected the dataset. X.H.Z., Z.Z., Y.H.L., and S.Z. designed and validated the models. Q.M.X., X.H.Z., Z.Z., and Y.H.L. analyzed the data and created the figures. Q.M.X., X.H.Z., Z.Z., Y.H.L, P.Y.X.. wrote the manuscript. X.L.L., Y.Z.Y., L.J.Z, O.G., and G.M.L. supervised the work. Q.M.X.and X.H.Z. are co-first authors of this paper.

## Ethics declarations

## Reference

1 WHO. Weekly Epidemiological and Operational updates October 2020. https://www.who.int/docs/default-source/coronaviruse/situation-reports/20201012-weekly-epi-update-9.pdf.

2 Vincent JL, Taccone FS. Understanding pathways to death in patients with COVID-19. Lancet Respir Med 2020; 8(5):430–432.

3 Kissler SM, Tedijianto C, Goldstein E, Yonatan HG, Lipsitch M. Projecting the transmission dynamics of SARS-CoV-2 through the postpandemic period. Science 2020; 368(6493): 860–868.

4 Harvard Business Review. We Need to Relocate ICU Patients Out of Covid-19 Hotspots. https://hbr.org/2020/06/we-need-to-relocate-icu-patients-out-of-covid-19-hotspots. Date: 2020. Date accessed: June 23, 2020.

5 BBC News. Coronavirus: Thousands of extra hospital beds and staff. https://www.bbc.com/news/uk-51989183. Date: 2020. Date accessed: March 21, 2020.

6 Chen C, Zhao B. Makeshift hospitals for COVID-19 patients: Where health-care workers and patients need sufficient ventilation for more protection. J. Hosp. Infect 2020; 105(1):98–99.

7 AP NEWS. Pentagon says it will give 5 million respirators, 2,000 ventilators to Health and Human Services for virus response. https://apnews.com/79e98812b5b1592a134803b00c8d88b0. xDate: 2020. Date accessed: March 17, 2020.

8 BBC News. Coronavirus: How can China build a hospital so quickly? https://www.bbc.com/news/world-asia-china-51245156. xDate: 2020. Date accessed: January 31, 2020.

9 Wu WH, Niu YY, Zhang CR, et al. Combined APACH II score and arterial blood lactate clearance rate to predict the prognosis of ARDS patients. Asian Pac J Trop Med 2012; 5:656–60.

10 Wang Y, Ju M, Chen C, et al. Neutrophil-to-lymphocyte ratio as a prognostic marker in acute respiratory distress syndrome patients: a retrospective study. J Thorac Dis 2018; 10:273–82.

11 Kumarasamy C, Sabarimurugan S, Madurantakam RM, et al. Prognostic significance of blood inflammatory biomarkers NLR, PLR, and LMR in cancer-A protocol for systematic review and meta-analysis. Medicine (Baltimore) 2019; 98:e14834.

12 Jiang J, Liu R, Yu X, et al. The neutrophil-lymphocyte count ratio as a diagnostic marker for bacteraemia: A systematic review and meta-analysis. Am J Emerg Med 2019; 37:1482–9.

13 China NHC. Diagnosis and treatment protocols of pneumonia caused by novel coronavirus (trial version 7). http://en.nhc.gov.cn/2020-03/29/c_78469.htm. Date: 2020. Date accessed: March 29, 2020.

14 Hanley JA, McNeil BJ. A method of comparing the areas under receiver operating characteristic curves derived from the same cases. Radiology 1983; 148.3: 839–843.

15 Saito T, Rehmsmeier M. The precision-recall plot is more informative than the ROC plot when evaluating binary classifiers on imbalanced datasets. PloS one 2015; 10.3: e0118432.

16 Cox DR. Regression models and life-tables. Journal of the Royal Statistical Society: Series B (Methodological) 1972; 34(2): 187–220.

17 Tibshirani R. “The lasso method for variable selection in the Cox model.” Statistics in Medicine, 1997; 16.4: 385–395.

18 Pölsterl S, Navab N, Katouzian A, Fast Training of Support Vector Machines for Survival Analysis. Machine Learning and Knowledge Discovery in Databases: European Conference. ECML PKDD 2015; Lecture Notes in Computer Science, 9285: 243–259.

19 Pölsterl S, Navab N, Katouzian A. An Efficient Training Algorithm for Kernel Survival Support Vector Machines. 4th Workshop on Machine Learning in Life Sciences 2016; 23 September, Riva del Garda, Italy. 1611.07054.

20 Truog RD, Mitchell C, Daley GQ. The Toughest Triage — Allocating Ventilators in a Pandemic. Engl J Med 2020; 382:1973–1975.

21 Emanuel EJ, Persad G, Upshur R, et al. Fair Allocation of Scarce Medical Resources in the Time of Covid-19. N Engl J Med 2020; 382:2049–2055.

22 Yu Q, Wang Y, Huang S, et al. Multicenter cohort study demonstrates more consolidation in upper lungs on initial CT increases the risk of adverse clinical outcome in COVID-19 patients. Theranostics 2020;10:5641–5648.

23 Chan JF, Yuan SF, Kok KH, et al. A familial cluster of pneumonia associated with the 2019 novel coronavirus indicating person-to-person transmission: a study of a family cluster. Lancet 2020; 395(10223): 514–523.

24 Li K, Wu J, Wu F, et al. The clinical and chest CT features associated with severe and critical COVID-19 pneumonia. Invest Radiol 2020; 55(6): 327–331.

25 Tabatabaei SMH, Talari H, Moghaddas F, Rajebi H. Computed Tomographic Features and Short-term Prognosis of Coronavirus Disease 2019 (COVID-19) Pneumonia: A Single-Center Study from Kashan, Iran. Radiology: Cardiothoracic Imaging 2020; 2(2).

26 Zhang Z, Liu XH, Shen J, et al. Clinically Applicable AI System for Accurate Diagnosis, Quantitative Measurements, and Prognosis of COVID-19 Pneumonia Using Computed Tomography. Cell 2020; 181(6): 1423-1433.e11.

27 Liang WH, Yao JH, Chen A, et al. Early triage of critically ill COVID-19 patients using deep learning. Nat Commun 2020; 11, 3543.

28 Cohen PA, Hall L, Johns JN, Rapoport AB. The early natural history of SARS-CoV-2 infection: clinical observations from an urban, ambulatory COVID-19 clinic. Mayo Clin Proc 2020; 95(6): 1124–1126.

29 Liang WH, Liang HG, Ou L, et al. Development and Validation of a Clinical Risk Score to Predict the Occurrence of Critical Illness in Hospitalized Patients With COVID-19. JAMA Intern Med 2020; doi:10.1001/jamainternmed.2020.2033.

30 Zhou Y, Fu B, Zheng X, et al. Aberrant pathogenic GM-CSF+ T cells and inflammatory CD14+CD16+ monocytes in severe pulmonary syndrome patients of a new coronavirus. bioRxiv 2020; https://www.biorxiv.org/content/10.1101/ 2020.02.12.945576v1.full.pdf.

31 Xu Z, Shi L, Wang Y, et al. Pathological findings of COVID-19 associated with acute respiratory distress syndrome. Lancet Respir Med 2020;8:420–2.

32 Zhou F, Yu T, Du R et al. Clinical course and risk factors for mortality of adult inpatients with COVID-19 in Wuhan, China: a retrospective cohort study. The Lancet. 2020; 395(10229): 1054–1062.

33 Yang X, Yu Y, Xu J, et al. Clinical course and outcomes of critically ill patients with SARS-CoV-2 pneumonia in Wuhan, China: a single-centered, retrospective, observational study. Lancet Respir Med 2020; 8:5, 475-481.

34 Chen N, Zhou M, Dong X, et al. Epidemiological and clinical characteristics of 99 cases of 2019 novel coronavirus pneumonia in Wuhan, China: a descriptive study. Lancet 2020; 395: 507–13.

35 Guo L, Wei D, Zhang X, et al. Clinical features predicting mortality risk in patients with viral pneumonia: The MuLBSTA Score. Front Microbiol 2019; 10:2752.

36 Wu C, Chen X, Cai Y, et al. Risk factors associated with acute respiratory distress syndrome and death in patients with coronavirus disease 2019 pneumonia in Wuhan, China. JAMA Intern Med 2020; 180(7):934–943.

