## Supplementary figures and images for "CT-based Rapid Triage of COVID-19 Patients: Risk Prediction and Progression Estimation of ICU Admission, Mechanical Ventilation, and Death of Hospitalized Patients"

### Supplement Figure 1

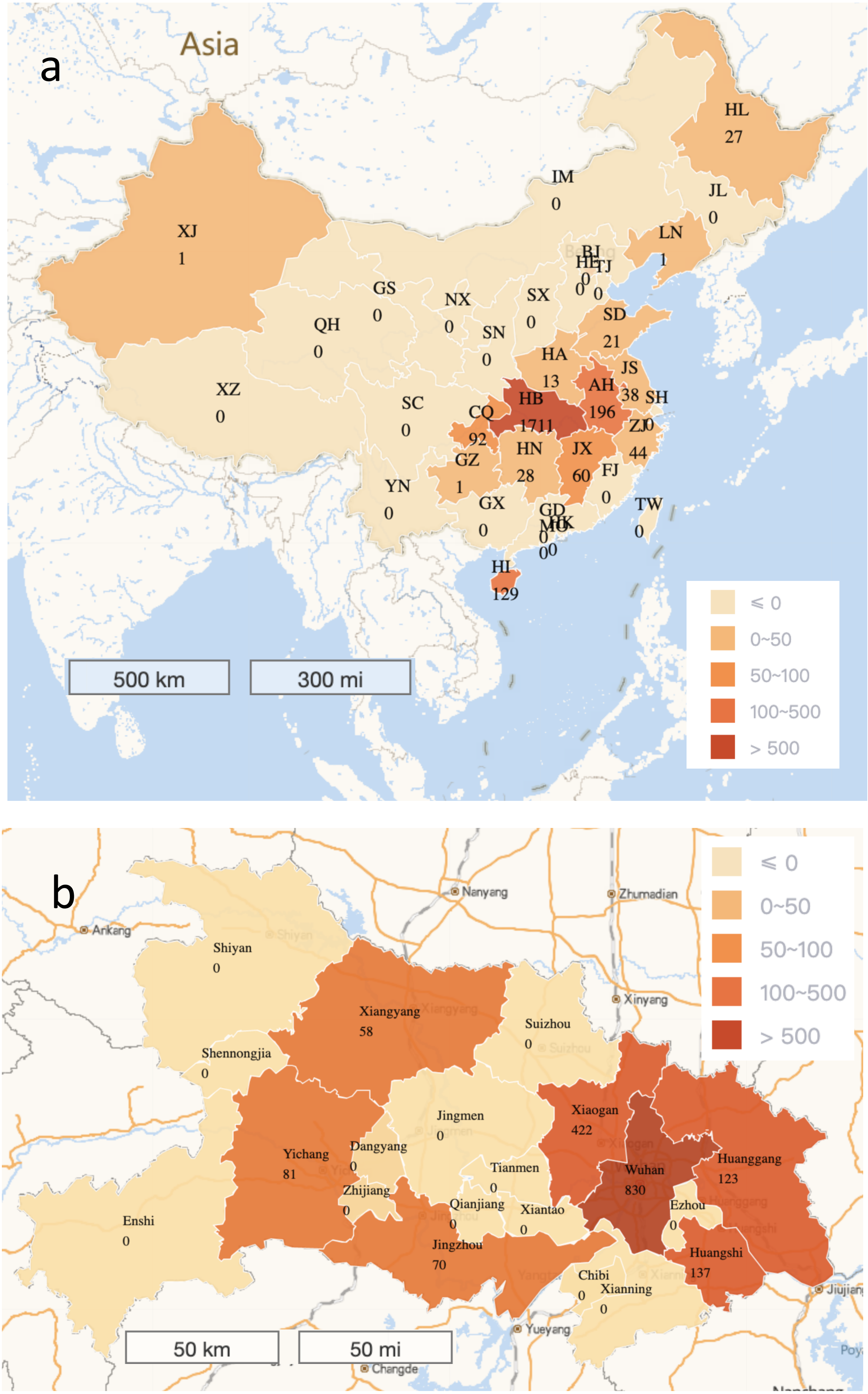

### Supplement Figure 2

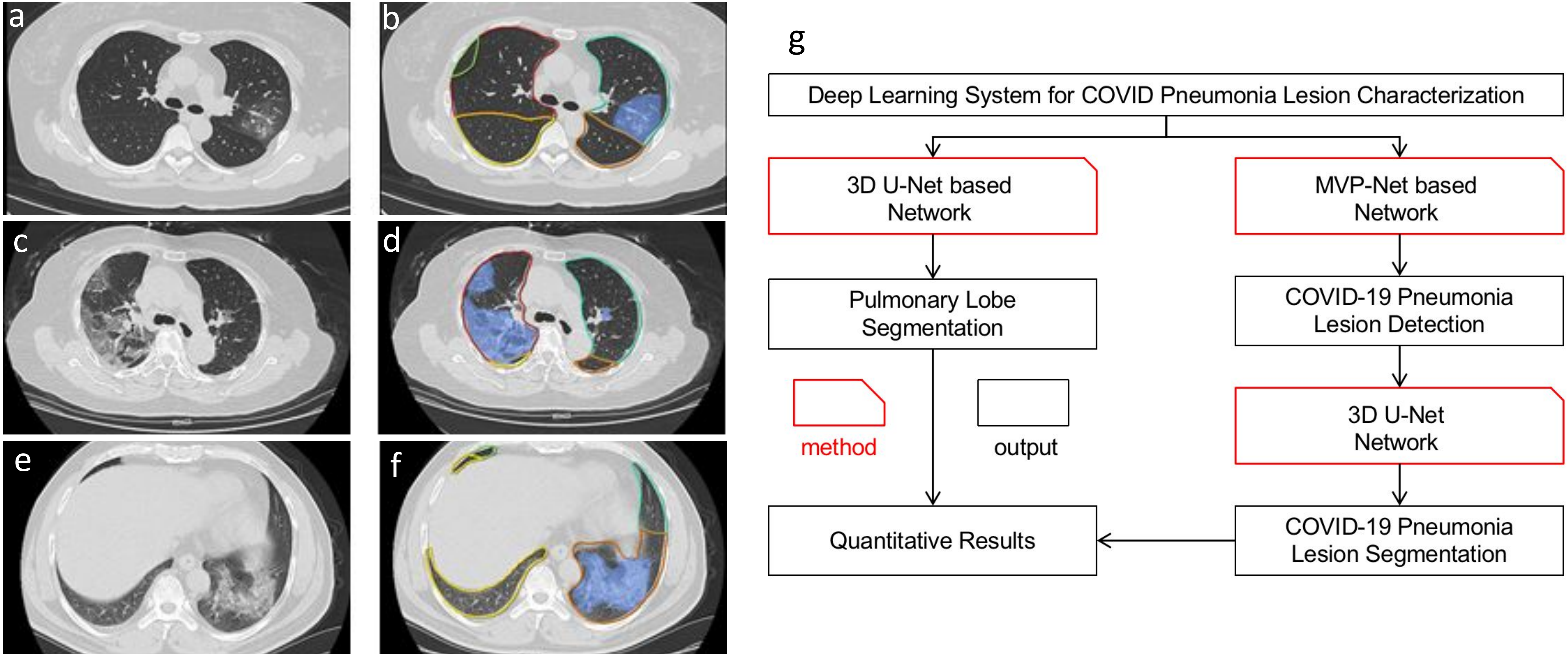

### Supplement Figure 3

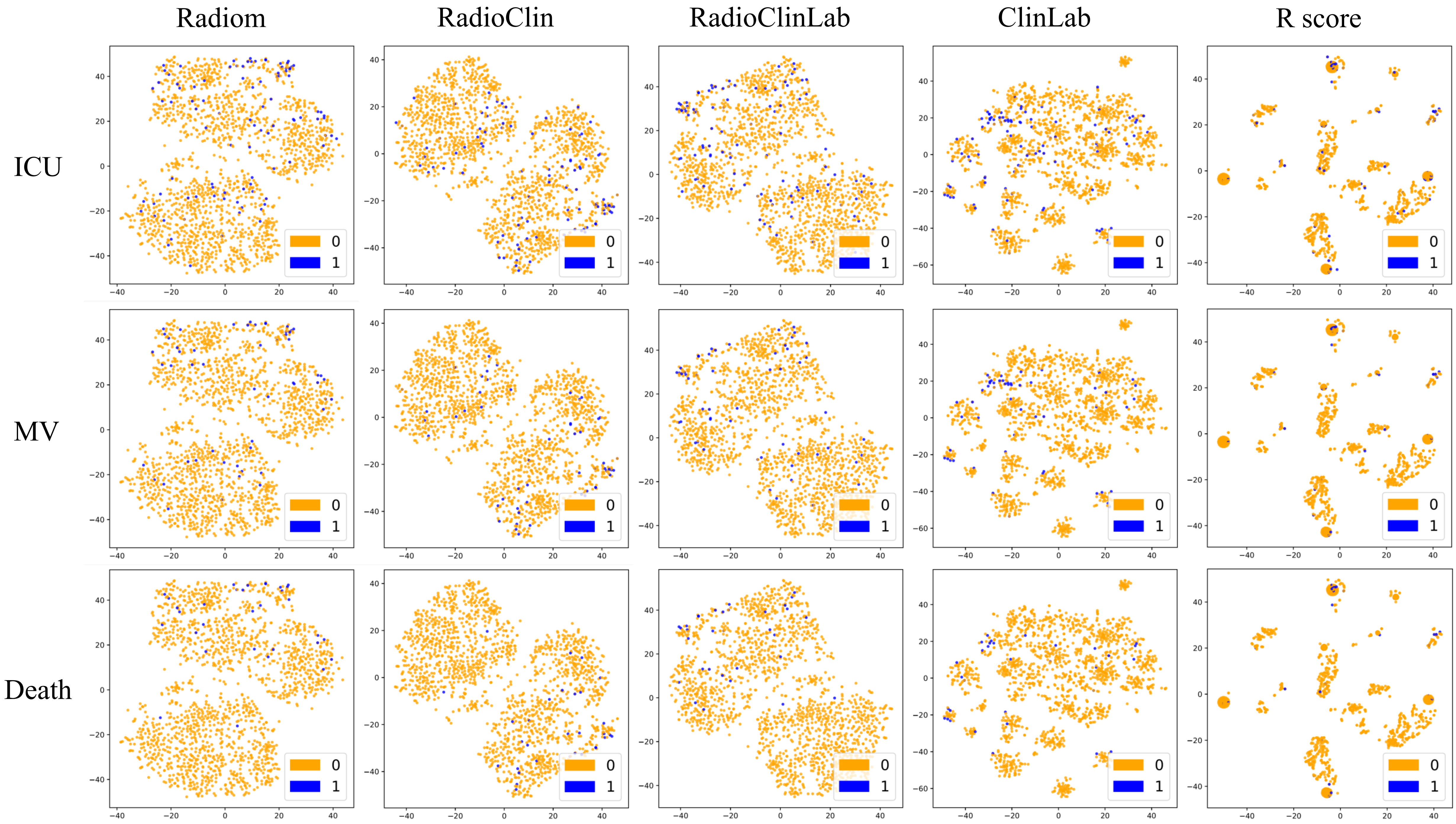

### Supplement Figure 4

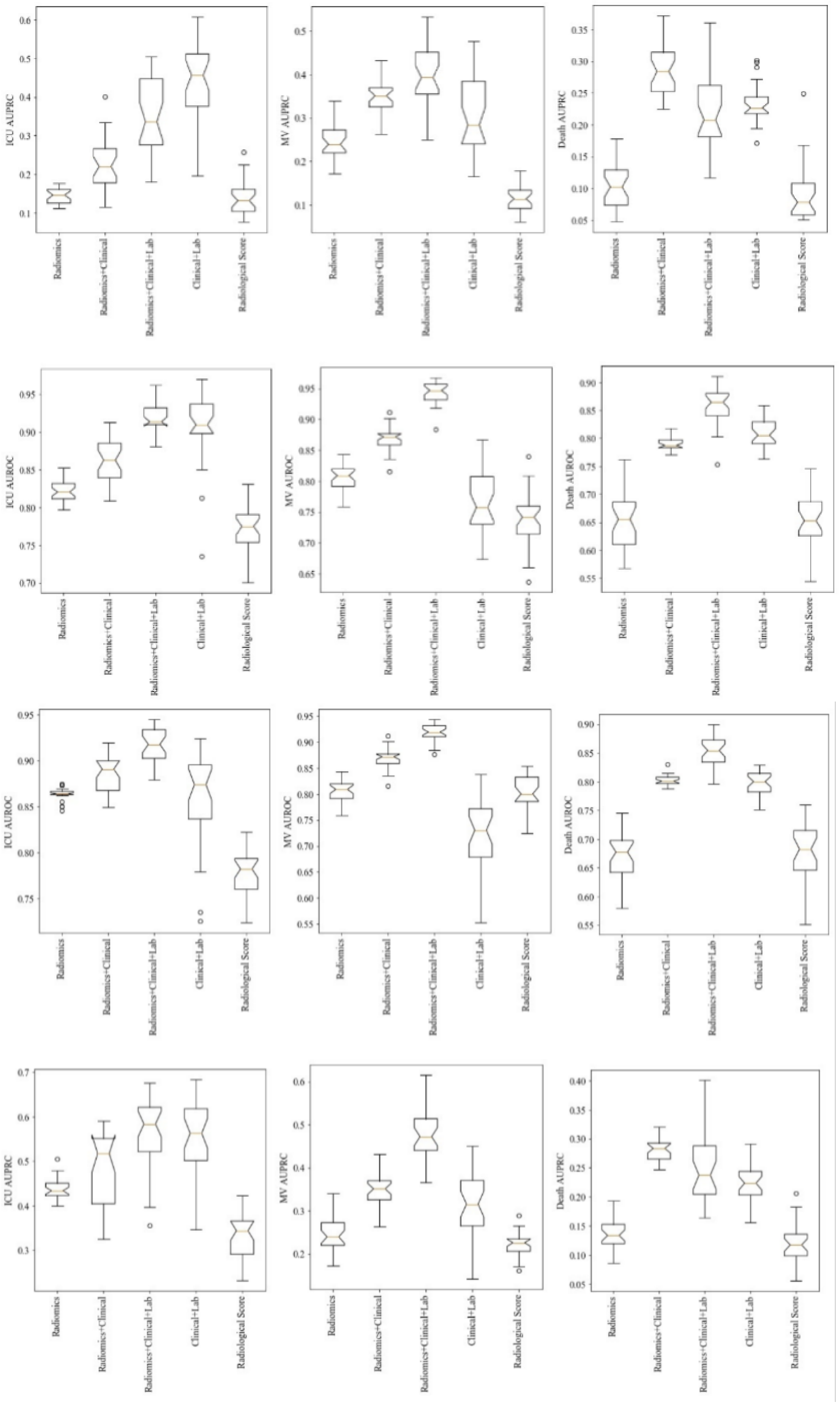

### Supplement Figure 5a

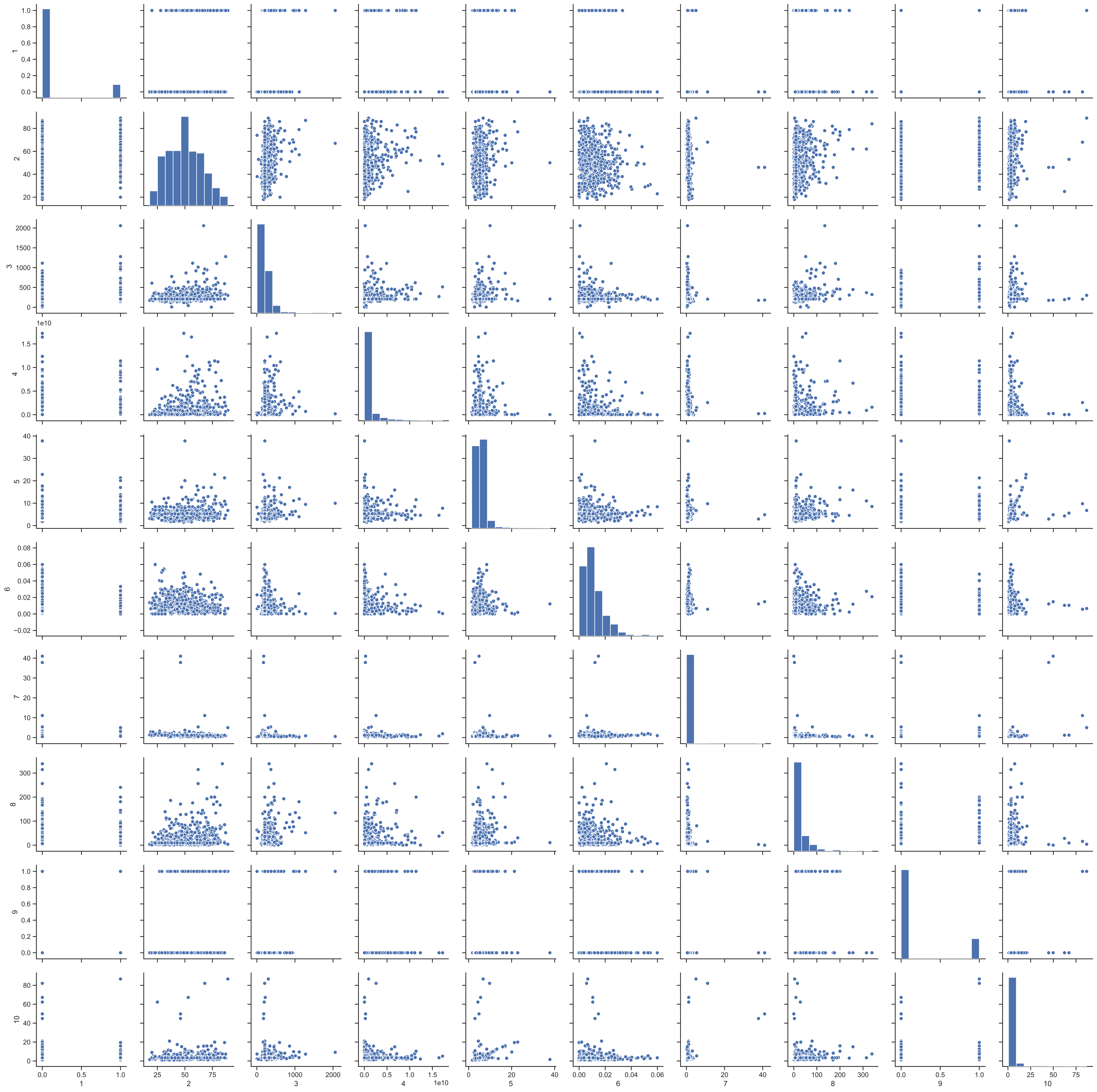

### Supplement Figure 5b

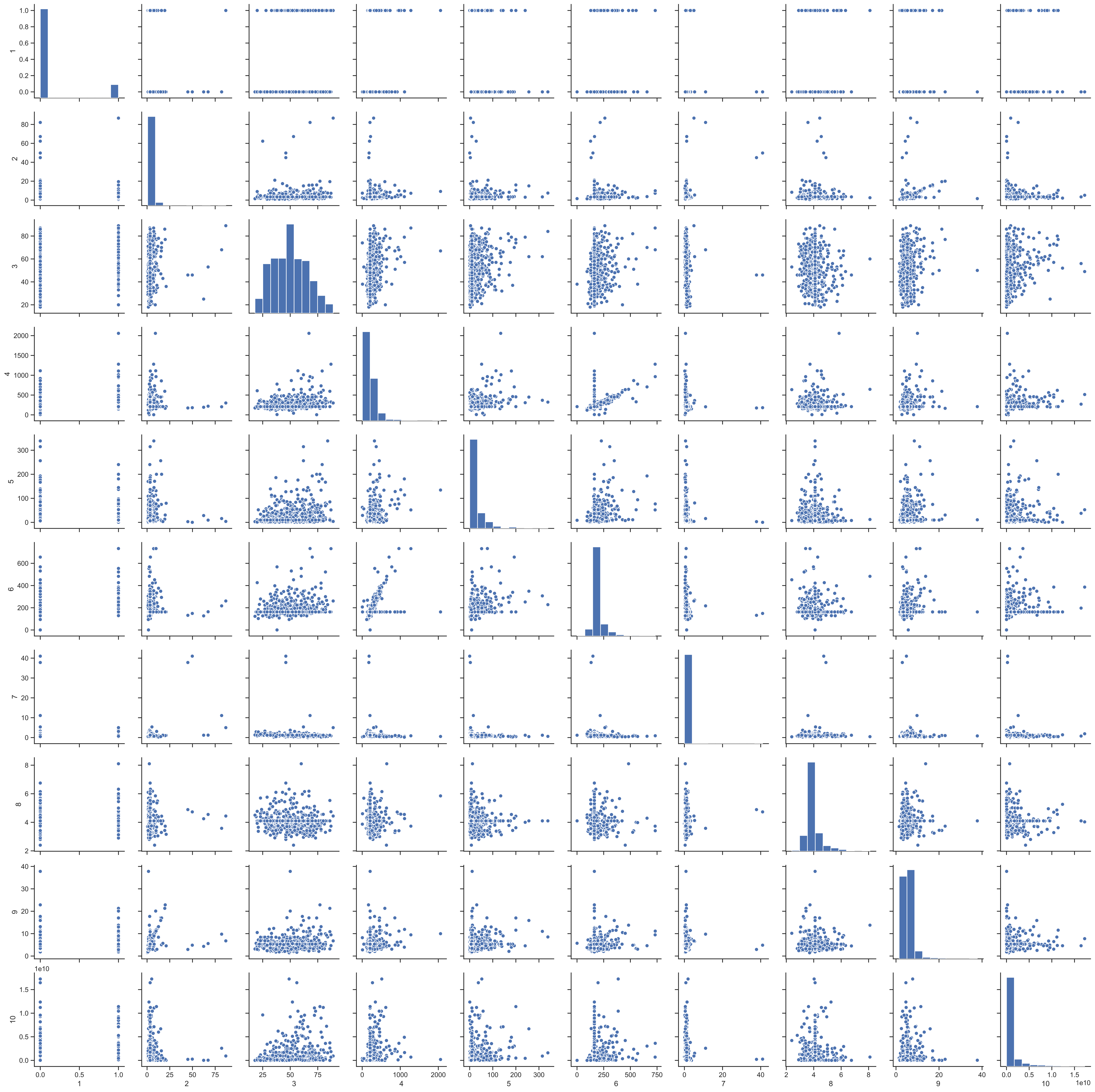

### Supplement Figure 6

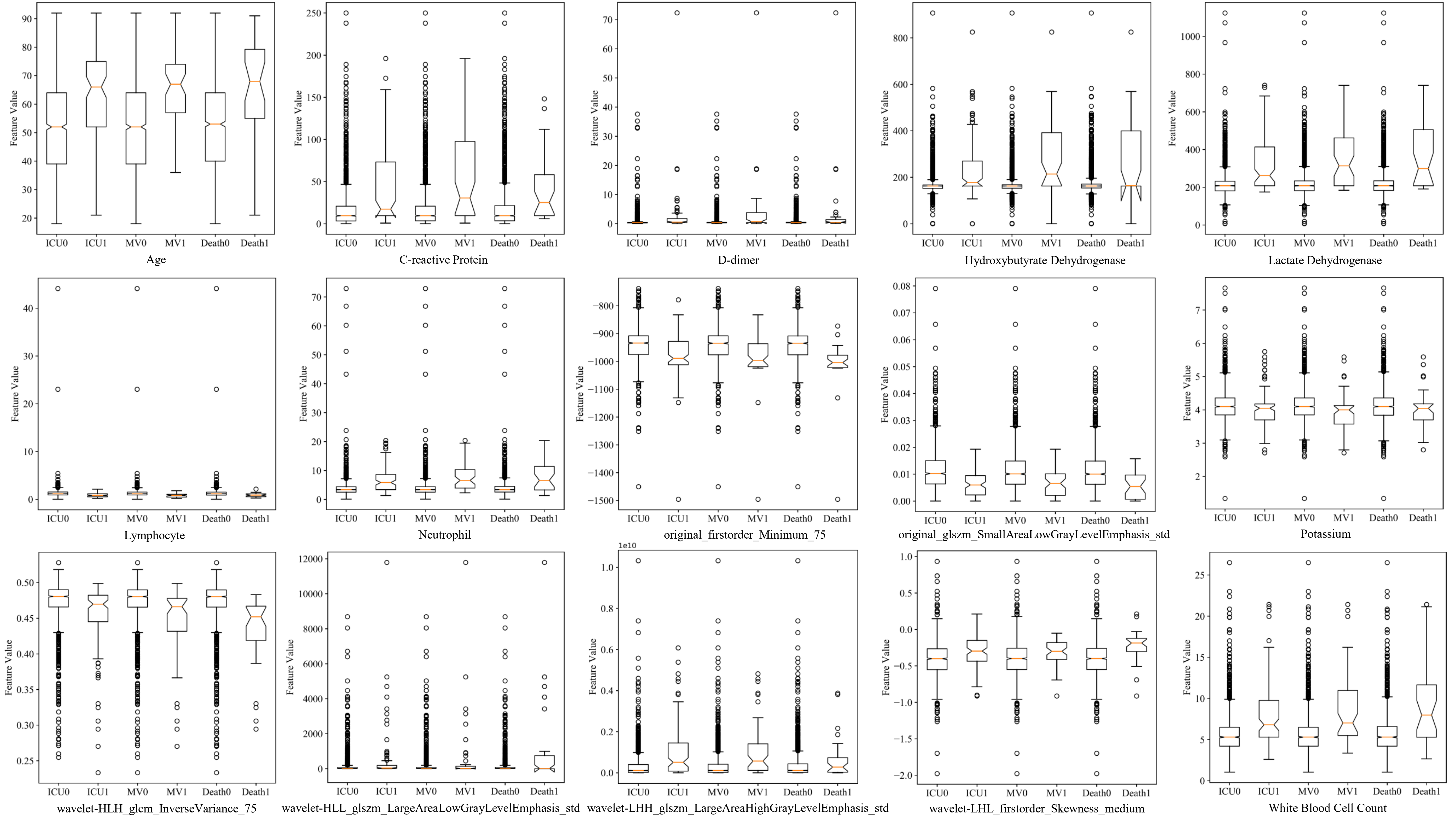

### Supplement Figure 7

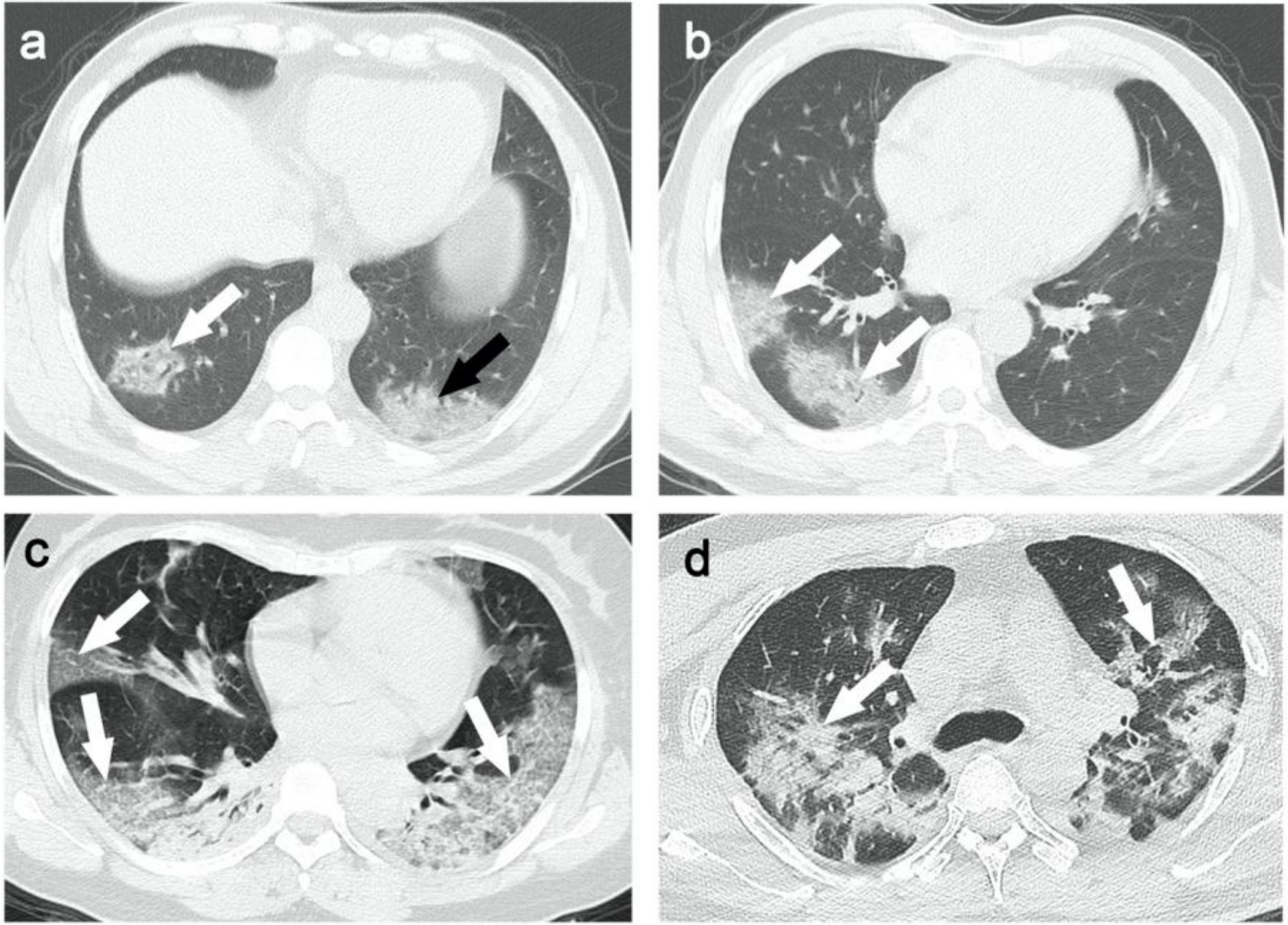

### Supplement Table 5c

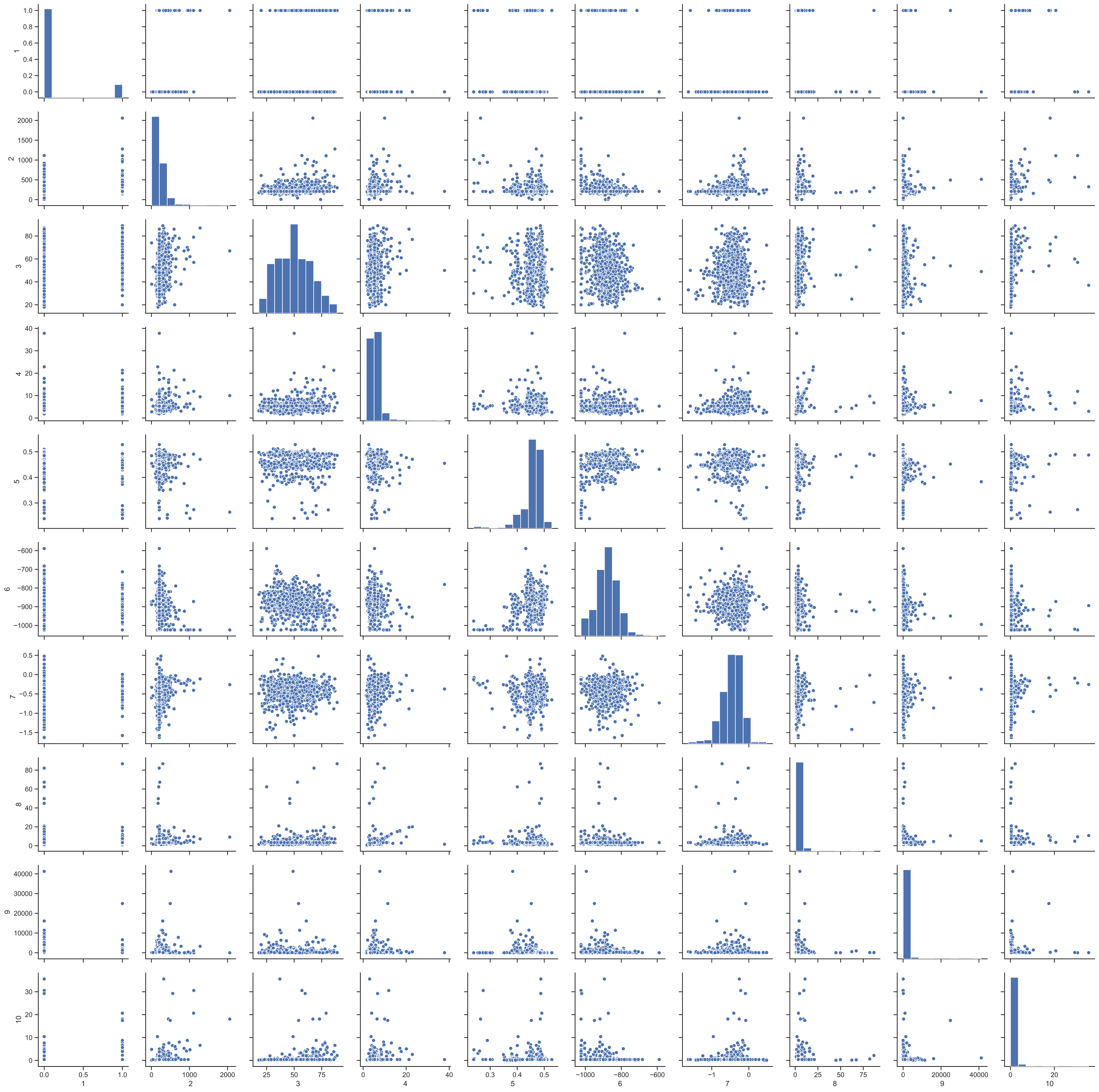
