## Supplement Tables for "CT-based Rapid Triage of COVID-19 Patients: Risk Prediction and Progression Estimation of ICU Admission, Mechanical Ventilation, and Death of Hospitalized Patients"

Table S1. Geographical distribution of COVID-19 patients in this study

| | | Cohort 1 | | | Cohort 2 | | |
|---|---|---|---|---|---|---|---|
| Province | Total (n = 2362) | Total (n = 1662) | Stable group | Adverse (ICU) group | Total (n = 700) | Stable group | Adverse (ICU) group |
| Hubei (HB) | 1721 | 1160 | 1080 | 80 | 561 | 513 | 48 |
| Wuhan | 830 | 830 | 782 | 48 | 0 | 0 | 0 |
| Huangshi | 137 | 137 | 123 | 14 | 0 | 0 | 0 |
| Huanggang | 123 | 123 | 110 | 13 | 0 | 0 | 0 |
| Jingzhou | 70 | 70 | 65 | 5 | 0 | 0 | 0 |
| Xiaogan | 422 | 0 | 0 | 0 | 422 | 389 | 33 |
| Yichang | 81 | 0 | 0 | 0 | 81 | 72 | 9 |
| Xiangyang | 58 | 0 | 0 | 0 | 58 | 52 | 6 |
| Anhui (AH) | 196 | 196 | 190 | 6 | 0 | 0 | 0 |
| Hainan (HI) | 129 | 129 | 124 | 5 | 0 | 0 | 0 |
| Chongqing (CQ) | 92 | 92 | 91 | 1 | 0 | 0 | 0 |
| Zhejiang (ZJ) | 44 | 44 | 44 | 0 | 0 | 0 | 0 |
| Jiangsu (JS) | 38 | 38 | 38 | 0 | 0 | 0 | 0 |
| Jiangxi (JX) | 51 | 0 | 0 | 0 | 51 | 51 | 0 |
| Hunan (HN) | 28 | 0 | 0 | 0 | 28 | 27 | 1 |
| Heilongjiang (HL) | 26 | 0 | 0 | 0 | 26 | 20 | 6 |
| Shandong (SD) | 21 | 0 | 0 | 0 | 21 | 20 | 1 |
| Henan (HA) | 13 | 0 | 0 | 0 | 13 | 13 | 0 |
| Xinjiang (XJ) | 1 | 1 | 1 | 0 | 0 | 0 | 0 |
| Guizhou (GZ) | 1 | 1 | 0 | 1 | 0 | 0 | 0 |
| Liaoning (LN) | 1 | 1 | 1 | 0 | 0 | 0 | 0 |

Note. The stable group refers to patients who discharged without any adverse outcome; the adverse group includes patients who developed adverse clinical outcomes and were admitted to the ICU (including patients required mechanical ventilation and those who died).

Table S2. Characteristics of patients in stable/adverse (ICU) groups, non-MV/MV groups, and discharge/death groups

| | Stable group (n = 2207) | Adverse (ICU) group (n = 155) | *P* value | Non-MV group (n = 2268) | MV group (n = 94) | *P* value | Discharge group (n = 2303) | Death group (n = 59) | *P* value |
|---|---|---|---|---|---|---|---|---|---|
| **Clinical feature** | | | | | | | | | |
| **Demographics** | | | | | | | | | |
| Age (years) | 50.802 ± 15.334 | 64.781 ± 14.179 | <.001 | 51.150 ± 15.501 | 65.590 ± 12.476 | <.001 | 51.320 ± 15.467 | 67.390 ± 14.643 | <.001 |
| Gender (male) | 1138 (51.5%) | 91 (58.7%) | .085 | 1176 (51.8%) | 53 (56.3%) | .389 | 1191 (51.7%) | 38 (64.4%) | .054 |
| **Comorbidity** | | | | | | | | | |
| Coronary heart disease | 135 (6.1%) | 37 (23.8%) | <.001 | 151 (6.6%) | 21 (22.3%) | <.001 | 152 (6.6%) | 20 (33.8%) | <.001 |
| Chronic liver disease | 81 (3.6%) | 1 (0.6%) | .047 | 81 (3.5%) | 1 (1.0%) | .257 | 81 (3.5%) | 1 (1.6%) | .721 |
| Chronic kidney disease | 20 (0.9%) | 9 (5.8%) | <.001 | 24 (1.0%) | 5 (5.3%) | .005 | 25 (1.0%) | 4 (6.7%) | .005 |
| COPD | 36 (1.6%) | 15 (9.6%) | <.001 | 41 (1.8%) | 10 (10.6%) | <.001 | 44 (1.9%) | 7 (11.8%) | <.001 |
| Diabetes | 224 (10.1%) | 37 (23.8%) | <.001 | 240 (10.5%) | 21 (22.3%) | <.001 | 245 (10.6%) | 16 (27.1%) | <.001 |
| Hypertension | 427 (19.3%) | 73 (47.0%) | <.001 | 455 (20.0%) | 45 (47.8%) | <.001 | 468 (20.3%) | 32 (54.2%) | <.001 |
| Carcinoma | 48 (2.1%) | 13 (8.3%) | <.001 | 51 (2.2%) | 10 (10.6%) | <.001 | 55 (2.3%) | 6 (10.1%) | .004 |
| **Clinical symptom** | | | | | | | | | |
| Fever | 1825 (82.6%) | 125 (80.6%) | .105 | 1873 (82.5%) | 77 (81.9%) | .274 | 1906 (82.7%) | 44 (74.5%) | .473 |
| Cough | 1540 (69.7%) | 111 (71.6%) | .529 | 1579 (69.6%) | 72 (76.5%) | .259 | 1608 (69.8%) | 43 (72.8%) | .891 |
| Myalgia | 517 (23.4%) | 36 (23.2%) | .163 | 529 (23.3%) | 24 (25.5%) | .148 | 543 (23.5%) | 10 (16.9%) | .039 |
| Fatigue | 887 (40.1%) | 65 (41.9%) | .130 | 910 (40.1%) | 42 (46.6%) | .111 | 930 (40.3%) | 22 (37.2%) | .002 |
| Headache | 179 (8.1%) | 12 (7.7%) | .022 | 183 (8.0%) | 8 (8.5%) | .084 | 184 (7.9%) | 7 (11.8%) | .023 |
| Nausea or vomiting | 104 (4.7%) | 12 (7.7%) | .093 | 111 (4.8%) | 5 (5.3%) | .319 | 109 (4.7%) | 7 (11.8%) | .522 |
| Diarrhea | 154 (6.9%) | 13 (8.3%) | .085 | 158 (6.9%) | 9 (9.5%) | .530 | 165 (7.1%) | 3 (3.3%) | .183 |
| Abdominal pain | 25 (1.1%) | 3 (1.9%) | .470 | 27 (1.1%) | 1 (1.0%) | .716 | 28 (1.2%) | 0 (0.0%) | .409 |

| | | | | | | | | | |
|---|---|---|---|---|---|---|---|---|---|
| Dyspnea | 322 (14.5%) | 125 (80.6%) | .728 | 346 (15.2%) | 57 (60.6%) | .303 | 369 (16.0%) | 34 (57.6%) | .353 |
| **Laboratory result** | | | | | | | | | |
| **Blood routine - no. (%)** | | | | | | | | | |
| WBC | | | <.001 | | | | | | <.001 |
| Increased | 97 (97 / 1893) | 37 (37 / 144) | | 106 (106 / 1945) | 28 (28 / 92) | <.001 | 110 (110 / 1980) | 24 (24 / 57) | |
| Decreased | 506 (506 / 1893) | 19 (19 / 144) | | 514 (514 / 1945) | 11 (11 / 92) | | 520 (520 / 1980) | 5 (5 / 57) | |
| Neutrophil | | | <.001 | | | <.001 | | | <.001 |
| Increased | 126 (126 / 1745) | 52 (52 / 140) | | 140 (140 / 1794) | 38 (38 / 91) | | 147 (147 / 1828) | 31 (31 / 57) | |
| Decreased | 300 (300 / 1745) | 9 (9 / 140) | | 306 (306 / 1794) | 3 (3 / 91) | | 307 (307 / 1828) | 2 (2 / 57) | |
| Lymphocyte | | | .698 | | | .376 | | | .608 |
| Increased | 10 (10 / 1801) | 1 (1 / 140) | | 11 (11 / 1850) | 38 (38 / 91) | | 10 (10 / 1885) | 1 (1 / 56) | |
| Decreased | 355 (355 / 1801) | 80 (80 / 140) | | 382 (382 / 1850) | 53 (53 / 91) | | 400 (400 / 1885) | 35 (35 / 56) | |
| Hemoglobin | | | <.001 | | | .005 | | | .170 |
| Increased | 140 (140 / 1770) | 5 (5 / 139) | | 141 (141 / 1822) | 4 (4 / 87) | | 141 (141 / 1856) | 4 (4 / 53) | |
| Decreased | 251 (251 / 1770) | 58 (58 / 139) | | 276 (276 / 1822) | 33 (33 / 87) | | 291 (291 / 1856) | 18 (18 / 53) | |
| Platelet | | | <.001 | | | .007 | | | .109 |
| Increased | 83 (83 / 1763) | 3 (3 / 138) | | 94 (94 / 1814) | 2 (2 / 87) | | 94 (94 / 1848) | 2 (2 / 53) | |
| Decreased | 93 (93 / 1763) | 27 (27 / 138) | | 96 (96 / 1814) | 14 (14 / 87) | | 102 (102 / 1848) | 8 (8 / 53) | |
| **Coagulation function** | | | | | | | | | |
| PT | | | <.001 | | | .001 | | | .038 |
| Increased | 414 (414 / 1344) | 58 (58 / 124) | | 432 (432 / 1389) | 40 (40 / 79) | | 448 (448 / 1422) | 24 (24 / 46) | |
| Decreased | 165 (165 / 1344) | 6 (6 /124) | | 169 (169 / 1389) | 2 (2 / 79) | | 169 (169 / 1422) | 2 (2 / 46) | |
| aPTT | | | <.001 | | | .421 | | | .523 |
| Increased | 240 (240 / 1273) | 27 (27 / 112) | | 248 (248 / 1314) | 19 (19 / 71) | | 254 (254 / 1346) | 13 (13 / 39) | |

| | | | | | | | | | |
|---|---|---|---|---|---|---|---|---|---|
| Decreased | 48 (48 / 1273) | 11 (11 / 112) | | 53 (53 / 1314) | 6 (6 / 71) | | 55 (55 / 1346) | 4 (4 / 39) | |
| D dimer | | | | | | | | | |
| Increased | 735 (735 / 1231) | 98 (98 / 111) | <.001 | 768 (768 / 1270) | 65 (65 / 72) | <.001 | 798 (798 / 1304) | 35 (35 / 38) | <.001 |
| **Infection-related biomarker** | | | | | | | | | |
| CRP | | | <.001 | | | <.001 | | | <.001 |
| Increased | 993 (993 / 1749) | 102 (102 / 129) | | 1028 (1028 / 1796) | 67 (67 / 82) | | 1054 (1054 / 1829) | 41 (41 / 49) | |
| **Blood biochemistry** | | | | | | | | | |
| Albumin | | | <.001 | | | <.001 | | | .103 |
| Increased | 59 (59 / 1653) | 1 (1 / 136) | | 59 (59 / 1703) | 1 (1 / 86) | | 59 (59 / 1740) | 1 (1 / 49) | |
| Decreased | 345 (345 / 1653) | 70 (70 / 136) | | 369 (369 / 1703) | 46 (46 / 86) | | 383 (383 / 1740) | 32 (32 / 49) | |
| ALT | | | <.001 | | | .001 | | | .165 |
| Increased | 406 (406 / 1596) | 50 (50 / 123) | | 425 (425 / 1641) | 31 (31 / 78) | | 440 (440 / 1675) | 16 (16 / 44) | |
| AST | | | .005 | | | .120 | | | .040 |
| Increased | 215 (215 / 1406) | 43 (43 / 126) | | 229 (229 / 1449) | 29 (29 / 83) | | 236 (236 / 1484) | 22 (22 / 48) | |
| Decreased | 144 (144 / 1406) | 11 (11 / 126) | | 145 (145 / 1449) | 10 (10 / 83) | | 150 (150 / 1484) | 5 (5 / 48) | |
| Total bilirubin | | | .139 | | | .235 | | | .610 |
| Increased | 209 (209 / 1527) | 42 (42 / 116) | | 222 (222 / 1568) | 29 (29 / 75) | | 232 (232 / 1601) | 19 (19 / 42) | |
| Decreased | 15 (15 / 1527) | 0 (0 / 116) | | 15 (15 / 1568) | 0 (0 / 75) | | 15 (15 / 1601) | 0 (0 / 42) | |
| Serum potassium | | | .520 | | | .251 | | | .642 |
| Increased | 113 (113 / 1558) | 13 (13 / 132) | | 118 (118 / 1605) | 8 (8 / 85) | | 120 (120 / 1639) | 6 (6 / 51) | |
| Decreased | 208 (208 / 1558) | 30 (30 / 132) | | 214 (214 / 1605) | 24 (24 / 85) | | 223 (223 / 1639) | 15 (15 / 51) | |
| Sodium | | | .001 | | | .018 | | | .036 |
| Increased | 60 (60 / 1505) | 30 (30 / 132) | | 70 (70 / 1551) | 20 (20 / 86) | | 76 (76 / 1587) | 14 (14 / 50) | |

| | | | | | | | | | | |
|---|---|---|---|---|---|---|---|---|---|---|
| | Decreased | 160 (160 / 1505) | 31 (31 / 132) | | 170 (170 / 1551) | 21 (21 / 86) | | 177 (177/ 1587) | 14 (14 / 50) | |
| Creatinine | | | | <.001 | | | .009 | | | <.001 |
| | Increased | 69 (69 / 1589) | 31 (31 / 129) | | 82 (82 / 1634) | 18 (18 / 84) | | 78 (78 / 1668) | 22 (22 / 50) | |
| | Decreased | 220 (220 / 1589) | 29 (29 / 129) | | 229 (229 / 1634) | 20 (20 / 84) | | 242 (242 / 1668) | 7 (7 / 50) | |
| CK | | | | .034 | | | .014 | | | <.001 |
| | Increased | 331 (331 / 1441) | 44 (44 / 121) | | 343 (343 / 1486) | 32 (32 / 76) | | 350 (350 / 1520) | 25 (25 / 42) | |
| | Decreased | 638 (638 / 1441) | 53 (53 / 121) | | 659 (659 / 1486) | 32 (32 / 76) | | 683 (683 / 1520) | 8 (8 / 42) | |
| LDH | | | | <.001 | | | <.001 | | | <.001 |
| | Increased | 480 (1416) | 95 (95 / 117) | | 512 (512 / 1455) | 63 (63 / 78) | | 538 (538 / 1491) | 37 (37 / 42) | |
| HBDH | | | | .262 | | | .397 | | | |
| | Increased | 488 (488 / 1132) | 67 (67 / 87) | | 505 (505 / 1157) | 50 (50 / 62) | | 524 (524 / 1180) | 31 (31 / 39) | .084 |
| | Decreased | 14 (14 / 1132) | 4 (4 / 87) | | 15 (15 / 1157) | 3 (3 / 62) | | 15 (15 / 1180) | 3 (3 / 39) | |

Note. The stable group refers to patients who discharged without any adverse outcome; the adverse group includes patients who developed adverse clinical outcomes and were admitted to the ICU (including patients required mechanical ventilation and those who died). p value is statistics of comparison between stable and adverse groups. The normal range refers to the criteria of each hospital. Increased means over the upper limit of the normal range and decreased means below the lower limit of the normal range. ± indicates mean ± standard deviation. Data in parentheses show percentage. COPD = Chronic obstructive lung disease; WBC = White blood cell; PT = Prothrombin time; aPTT = Activated partial thromboplastin time; CRP = C-reactive protein; ALT = Alanine aminotransferase; AST = Aspartate aminotransferase; CK = Creatine kinase; LDH = Lactate dehydrogenase; HBDH = α-Hydroxybutyrate dehydrogenase.

**Table S3. Statistical significance test of top ten important feature values of positive cases between Cohort 1 and Cohort 2**

| ICU | | |
|---|---|---|
| **Feature** | **Z_score** | **p_value** |
| Dyspnea | -1.613 | 0.107 |
| Age | -1.268 | 0.207 |
| LDH | -4.122 | <0.001 |
| wavelet-LHH_glszm_LargeAreaHighGrayLevelEmphasis_std | -3.206 | 0.001 |
| WBC | 1.732 | 0.083 |
| original_glszm_SmallAreaLowGrayLevelEmphasis_std | -0.359 | 0.719 |
| Lymphocyte | -0.945 | 0.345 |
| C-reactive Protein | -0.258 | 0.796 |
| Hypertension | -1.021 | 0.307 |
| Neutrophil | -1.013 | 0.311 |
| **MV** | | |
| **Feature** | **Z_score** | **p_value** |
| Dyspnea | -0.042 | 0.967 |
| Neutrophil | 0.837 | 0.403 |
| Age | 0.129 | 0.898 |
| LDH | -3.017 | 0.003 |
| C-reactive Protein | 0.284 | 0.776 |
| HBDH | 0.269 | 0.788 |
| Lymphocyte | 1.211 | 0.226 |
| Potassium | -0.214 | 0.830 |
| WBC | 1.152 | 0.249 |
| wavelet-LHH_glszm_LargeAreaHighGrayLevelEmphasis_std | -2.328 | 0.020 |
| **Death** | | |
| **Feature** | **Z_score** | **p_value** |
| Dyspnea | -0.350 | 0.726 |
| LDH | -0.682 | 0.495 |
| Age | -0.470 | 0.640 |
| WBC | 0.104 | 0.917 |
| wavelet-HLH_glcm_InverseVariance_75 | -0.696 | 0.486 |
| original_firstorder_Minimum_75 | -4.393 | <0.001 |
| wavelet-LHL_firstorder_Skewness_medium | 1.793 | 0.073 |
| Neutrophil | -0.407 | 0.684 |
| wavelet-HLL_glszm_LargeAreaLowGrayLevelEmphasis_std | -2.126 | 0.033 |
| D-dimer | -1.252 | 0.211 |

Table S4. Performance and algorithms of the optimal models of each data type for the prediction of ICU, MV, and death on the three cohorts

**ICU**

| Data | Feature Engineering | Model | Cohort 1 (n=1662) AUROC | Cohort 1 (n=1662) ACC | Cohort 1 (n=1662) AUPRC | Cohort 2 (n = 700) AUROC | Cohort 2 (n = 700) ACC | Cohort 2 (n = 700) AUPRC | Cohort 3 (n = 662) AUROC | Cohort 3 (n = 662) ACC | Cohort 3 (n = 662) AUPRC |
|---|---|---|---|---|---|---|---|---|---|---|---|
| Radiom | SMOTEENN, LASSO C=50 | MLP | 0.732 | 0.780 | 0.261 | 0.875 | 0.901 | 0.482 | 0.853 | 0.914 | 0.183 |
| RadioClin | SMOTEENN, LASSO C=0.5 | LightGBM | 0.836 | 0.826 | 0.383 | 0.919 | 0.923 | 0.554 | 0.919 | 0.946 | 0.332 |
| RadioClinLab | SMOTEENN, LASSO C=1 | LightGBM | 0.837 | 0.824 | 0.307 | 0.944 | 0.924 | 0.665 | 0.948 | 0.932 | 0.471 |
| ClinLab | SMOTEENN, LASSO C=0.2 | LR | 0.876 | 0.784 | 0.335 | 0.911 | 0.764 | 0.626 | 0.958 | 0.759 | 0.539 |
| R-score | / | LR | 0.600 | 0.939 | 0.096 | 0.823 | 0.916 | 0.444 | 0.813 | 0.968 | 0.167 |

**MV**

| Data | Feature Engineering | Model | Cohort 1 (n=1662) AUROC | Cohort 1 (n=1662) ACC | Cohort 1 (n=1662) AUPRC | Cohort 2 (n = 700) AUROC | Cohort 2 (n = 700) ACC | Cohort 2 (n = 700) AUPRC | Cohort 3 (n = 662) AUROC | Cohort 3 (n = 662) ACC | Cohort 3 (n = 662) AUPRC |
|---|---|---|---|---|---|---|---|---|---|---|---|
| Radiom | SMOTEENN 1:3 | LightGBM | 0.823 | 0.954 | 0.307 | 0.799 | 0.946 | 0.247 | 0.753 | 0.968 | 0.154 |
| RadioClin | SMOTEENN 1:3 | LightGBM | 0.836 | 0.826 | 0.383 | 0.881 | 0.946 | 0.335 | 0.874 | 0.967 | 0.225 |
| RadioClinLab | SMOTEENN, LASSO C=1 | LightGBM | 0.850 | 0.970 | 0.420 | 0.942 | 0.949 | 0.551 | 0.955 | 0.967 | 0.425 |
| ClinLab | SMOTEENN 1:3 | MLP | 0.876 | 0.784 | 0.335 | 0.816 | 0.927 | 0.451 | 0.814 | 0.953 | 0.387 |
| R-score | / | LR | 0.607 | 0.967 | 0.065 | 0.829 | 0.944 | 0.251 | 0.670 | 0.970 | 0.105 |

**Death**

| Data | Feature Engineering | Model | Cohort 1 (n=1662) AUROC | Cohort 1 (n=1662) ACC | Cohort 1 (n=1662) AUPRC | Cohort 2 (n = 700) AUROC | Cohort 2 (n = 700) ACC | Cohort 2 (n = 700) AUPRC | Cohort 3 (n = 662) AUROC | Cohort 3 (n = 662) ACC | Cohort 3 (n = 662) AUPRC |
|---|---|---|---|---|---|---|---|---|---|---|---|
| Radiom | SMOTEENN, LASSO C=1 | RF | 0.881 | 0.970 | 0.300 | 0.687 | 0.960 | 0.192 | 0.680 | 0.968 | 0.123 |
| RadioClin | SMOTEENN, FPR, F-Classif | LR | 0.948 | 0.960 | 0.395 | 0.802 | 0.950 | 0.276 | 0.788 | 0.965 | 0.298 |
| RadioClinLab | SMOTEENN, LASSO C=1 | SVM | 0.826 | 0.984 | 0.417 | 0.860 | 0.963 | 0.346 | 0.882 | 0.973 | 0.337 |
| ClinLab | SMOTEENN, LASSO C=30 | MLP | 0.838 | 0.968 | 0.121 | 0.769 | 0.941 | 0.164 | 0.805 | 0.964 | 0.206 |
| R-score | / | LR | 0.704 | 0.979 | 0.056 | 0.694 | 0.960 | 0.119 | 0.670 | 0.970 | 0.105 |

Table S5. Statistical significance of the bootstrapping results of different data modalities in Cohort 2 and Cohort 3

| ICU | | | | | | | | | |
|---|---|---|---|---|---|---|---|---|---|
| Model comparison | | Cohort 2 | | | | Cohort 3 | | | |
| model 1 | model 2 | Z_AUROC | p-value | Z_AUPRC | p-value | Z_AUROC | p-value | Z_AUPRC | p-value |
| Radiom | R-score | 16.624 | <0.001 | 10.089 | <0.001 | 7.950 | <0.001 | 0.866 | 0.197 |
| RadioClin | Radiom | 5.591 | <0.001 | 2.538 | 0.008 | 6.545 | <0.001 | 6.147 | <0.001 |
| RadioClinLab | RadioClin | 7.834 | <0.001 | 5.17 | <0.001 | 7.994 | <0.001 | 4.969 | <0.001 |
| RadioClinLab | ClinLab | 5.702 | <0.001 | 0.816 | 0.211 | 1.244 | 0.112 | -3.834 | 0.001 |
| **MV** | | | | | | | | | |
| Model comparison | | Cohort 2 | | | | Cohort 3 | | | |
| model 1 | model 2 | Z_AUROC | p-value | Z_AUPRC | p-value | Z_AUROC | p-value | Z_AUPRC | p-value |
| Radiom | R-score | 0.254 | 0.401 | 2.947 | 0.003 | 7.834 | <0.001 | 15.648 | <0.001 |
| RadioClin | Radiom | 12.652 | <0.001 | 10.023 | <0.001 | 12.652 | <0.001 | 10.023 | <0.001 |
| RadioClinLab | RadioClin | 9.184 | <0.001 | 8.711 | <0.001 | 15.687 | <0.001 | 2.866 | 0.004 |
| RadioClinLab | ClinLab | 15.186 | <0.001 | 9.229 | <0.001 | 18.132 | <0.001 | 4.218 | <0.001 |
| **Death** | | | | | | | | | |
| Model comparison | | Cohort 2 | | | | Cohort 3 | | | |
| model 1 | model 2 | Z_AUROC | p-value | Z_AUPRC | p-value | Z_AUROC | p-value | Z_AUPRC | p-value |
| Radiom | R-score | -0.869 | 0.146 | 2.119 | 0.021 | 0.161 | 0.436 | 1.169 | 0.126 |
| RadioClin | Radiom | 19.518 | <0.001 | 25.452 | <0.001 | 14.206 | <0.001 | 20.231 | <0.001 |
| RadioClinLab | RadioClin | 11.486 | <0.001 | -3.654 | 0.001 | 9.956 | <0.001 | -5.646 | <0.001 |
| RadioClinLab | ClinLab | 9.217 | <0.001 | 2.106 | 0.022 | 5.336 | <0.001 | -0.969 | 0.170 |

**Table S6. The feature weight of traditional CT-based features on three outcome prediction tasks.**

| Feature | ICU | MV | Death |
|---|---|---|---|
| **Lesion distribution** | | | |
| Subpleural | -0.070 | -0.114 | 0.166 |
| Diffuse | 0.177 | 0.290 | 0.393 |
| **Lesion morphology** | | | |
| Round | -0.026 | -0.230 | -0.367 |
| Other | -0.023 | 0 | -0.086 |
| **Main sign** | | | |
| GGO | 0.137 | 0.141 | 0 |
| Pure consolidation | 0.137 | 0.156 | 0.055 |
| GGO with consolidation | 0.048 | 0.097 | 0.113 |
| ILD | -0.057 | 0 | 0 |
| Crazy-paving pattern | 0 | 0.098 | 0.117 |
| **Other abnormality** | | | |
| Pleural effusion | 0.224 | 0.205 | 0.150 |
| **Number of lesions in each lobe** | | | |
| RUL | 0 | 0.036 | -0.005 |
| RML | 0.134 | 0 | 0 |
| RLL | 0 | 0 | 0 |
| LUL | 0.357 | 0.292 | 0.131 |
| LLL | 0.014 | 0.013 | 0 |
| **Lesion count** | | | |
| Single | -0.007 | 0 | 0 |
| Multiple | 0 | 0 | -0.227 |

Note. GGO = the presence of pure ground-glass opacity; ILD = interstitial lung disease ILD; RRL = right upper lobe; RML = right middle lobe; RLL = right lower lobe; LUL = left upper lobe; LLL = left lower lobe

**Table S7. The statistical significance of the difference between negative and positive cases on all of the ten most important features in three outcome prediction tasks on Cohort 1.**

| Feature | Outcome | Top 10 Feature | Group 1 | Group 2 | Z score | p_value |
|---|---|---|---|---|---|---|
| wavelet-LHH_glszm_LargeAreaHighGrayLevelEmphasis_std | ICU | T | ICU1 | ICU0 | 5.710 | <0.001 |
| | MV | T | MV1 | MV0 | 4.897 | <0.001 |
| | Death | F | Death1 | Death0 | 1.667 | 0.096 |
| original_glszm_SmallAreaLowGrayLevelEmphasis_std | ICU | T | ICU1 | ICU0 | -6.751 | <0.001 |
| | MV | F | MV1 | MV0 | -4.622 | <0.001 |
| | Death | F | Death1 | Death0 | -4.267 | <0.001 |
| LDH | ICU | T | ICU1 | ICU0 | 7.522 | <0.001 |
| | MV | T | MV1 | MV0 | 6.766 | <0.001 |
| | Death | T | Death1 | Death0 | 5.335 | <0.001 |
| Age | ICU | T | ICU1 | ICU0 | 7.115 | <0.001 |
| | MV | T | MV1 | MV0 | 6.348 | <0.001 |
| | Death | T | Death1 | Death0 | 4.808 | <0.001 |
| WBC | ICU | T | ICU1 | ICU0 | 5.945 | <0.001 |
| | MV | T | MV1 | MV0 | 5.866 | <0.001 |
| | Death | T | Death1 | Death0 | 3.957 | <0.001 |
| Lymphocyte | ICU | T | ICU1 | ICU0 | -7.456 | <0.001 |
| | MV | T | MV1 | MV0 | -6.549 | <0.001 |
| | Death | F | Death1 | Death0 | -4.181 | <0.001 |
| Potassium | ICU | F | ICU1 | ICU0 | -2.359 | 0.018 |
| | MV | T | MV1 | MV0 | -2.842 | 0.004 |
| | Death | F | Death1 | Death0 | -1.325 | 0.185 |
| C-reactive Protein | ICU | T | ICU1 | ICU0 | 6.414 | <0.001 |
| | MV | T | MV1 | MV0 | 6.362 | <0.001 |
| | Death | F | Death1 | Death0 | 4.691 | <0.001 |
| Neutrophil | ICU | T | ICU1 | ICU0 | 7.341 | <0.001 |
| | MV | T | MV1 | MV0 | 7.680 | <0.001 |
| | Death | T | Death1 | Death0 | 4.418 | <0.001 |
| HBDH | ICU | F | ICU1 | ICU0 | 6.008 | <0.001 |
| | MV | T | MV1 | MV0 | 5.641 | <0.001 |
| | Death | F | Death1 | Death0 | 3.057 | 0.002 |
| wavelet-HLH_glcm_InverseVariance_75 | ICU | F | ICU1 | ICU0 | -5.085 | <0.001 |
| | MV | F | MV1 | MV0 | -5.156 | <0.001 |
| | Death | T | Death1 | Death0 | -6.302 | <0.001 |
| original_firstorder_Minimum_75 | ICU | F | ICU1 | ICU0 | -5.634 | <0.001 |
| | MV | F | MV1 | MV0 | -5.447 | <0.001 |
| | Death | T | Death1 | Death0 | -6.211 | <0.001 |
| wavelet-LHL_firstorder_Skewness_medium | ICU | F | ICU1 | ICU0 | 4.555 | <0.001 |
| | MV | F | MV1 | MV0 | 3.534 | <0.001 |
| | Death | T | Death1 | Death0 | 4.889 | <0.001 |
| wavelet-HLL_glszm_LargeAreaLowGrayLevelEmphasis_std | ICU | F | ICU1 | ICU0 | 0.941 | 0.346 |
| | MV | F | MV1 | MV0 | -0.584 | 0.559 |
| | Death | T | Death1 | Death0 | -0.521 | 0.602 |
| D-dimer | ICU | F | ICU1 | ICU0 | 5.909 | <0.001 |
| | MV | F | MV1 | MV0 | 5.581 | <0.001 |
| | Death | T | Death1 | Death0 | 3.249 | 0.001 |

| | | | | | | |
|---|---|---|---|---|---|---|
| Dyspnea | ICU | T | ICU1 | ICU0 | 7.094 | <0.001 |
| | MV | T | MV1 | MV0 | 7.572 | <0.001 |
| | Death | T | Death1 | Death0 | 5.099 | <0.001 |
| Hypertension | ICU | T | ICU1 | ICU0 | 5.117 | <0.001 |
| | MV | F | MV1 | MV0 | 3.972 | <0.001 |
| | Death | F | Death1 | Death0 | 2.942 | 0.003 |

Table S8. The statistical significance of the difference between negative and positive cases on all of the ten most important features in three outcome prediction tasks on Cohort 2

| Feature | Outcome | Top 10 Feature | Group 1 | Group 2 | Z score | p_value |
|---|---|---|---|---|---|---|
| wavelet-LHH_glszm_LargeAreaHighGrayLevelEmphasis_std | ICU | T | ICU1 | ICU0 | 5.940 | <0.001 |
| | MV | T | MV1 | MV0 | 4.801 | <0.001 |
| | Death | F | Death1 | Death0 | 2.825 | 0.005 |
| original_glszm_SmallAreaLowGrayLevelEmphasis_std | ICU | T | ICU1 | ICU0 | -4.333 | <0.001 |
| | MV | F | MV1 | MV0 | -2.982 | 0.003 |
| | Death | F | Death1 | Death0 | -2.151 | 0.032 |
| LDH | ICU | T | ICU1 | ICU0 | 8.774 | <0.001 |
| | MV | T | MV1 | MV0 | 7.535 | <0.001 |
| | Death | T | Death1 | Death0 | 4.795 | <0.001 |
| Age | ICU | T | ICU1 | ICU0 | 8.402 | <0.001 |
| | MV | T | MV1 | MV0 | 6.523 | <0.001 |
| | Death | T | Death1 | Death0 | 5.766 | <0.001 |
| WBC | ICU | T | ICU1 | ICU0 | 3.239 | 0.001 |
| | MV | T | MV1 | MV0 | 3.471 | 0.001 |
| | Death | T | Death1 | Death0 | 3.828 | <0.001 |
| Lymphocyte | ICU | T | ICU1 | ICU0 | -6.192 | <0.001 |
| | MV | T | MV1 | MV0 | -6.635 | <0.001 |
| | Death | F | Death1 | Death0 | -6.077 | <0.001 |
| Potassium | ICU | F | ICU1 | ICU0 | -0.554 | 0.580 |
| | MV | T | MV1 | MV0 | -1.059 | 0.290 |
| | Death | F | Death1 | Death0 | -2.160 | 0.031 |
| C-reactive Protein | ICU | T | ICU1 | ICU0 | 3.193 | 0.001 |
| | MV | T | MV1 | MV0 | 3.423 | 0.001 |
| | Death | F | Death1 | Death0 | 2.661 | 0.008 |
| Neutrophil | ICU | T | ICU1 | ICU0 | 4.877 | <0.001 |
| | MV | T | MV1 | MV0 | 5.534 | <0.001 |
| | Death | T | Death1 | Death0 | 5.481 | <0.001 |
| HBDH | ICU | F | ICU1 | ICU0 | 2.453 | 0.014 |
| | MV | T | MV1 | MV0 | 3.315 | 0.001 |
| | Death | F | Death1 | Death0 | 4.490 | <0.001 |
| wavelet-HLH_glcm_InverseVariance_75 | ICU | F | ICU1 | ICU0 | -2.586 | 0.010 |
| | MV | F | MV1 | MV0 | -2.236 | 0.025 |
| | Death | T | Death1 | Death0 | -1.878 | 0.060 |
| original_firstorder_Minimum_75 | ICU | F | ICU1 | ICU0 | -6.993 | <0.001 |
| | MV | F | MV1 | MV0 | -5.670 | <0.001 |
| | Death | T | Death1 | Death0 | -3.972 | <0.001 |

| | | | | | | |
|---|---|---|---|---|---|---|
| wavelet-LHL_firstorder_Skewness_medium | ICU | F | ICU1 | ICU0 | 5.687 | <0.001 |
| | MV | F | MV1 | MV0 | 3.528 | <0.001 |
| | Death | T | Death1 | Death0 | 2.346 | 0.019 |
| wavelet-HLL_glszm_LargeAreaLowGrayLevelEmphasis_std | ICU | F | ICU1 | ICU0 | 1.051 | 0.293 |
| | MV | F | MV1 | MV0 | 1.536 | 0.124 |
| | Death | T | Death1 | Death0 | 2.779 | 0.005 |
| D-dimer | ICU | F | ICU1 | ICU0 | 8.716 | <0.001 |
| | MV | F | MV1 | MV0 | 7.106 | <0.001 |
| | Death | T | Death1 | Death0 | 5.154 | <0.001 |
| Dyspnea | ICU | T | ICU1 | ICU0 | 11.667 | <0.001 |
| | MV | T | MV1 | MV0 | 9.232 | <0.001 |
| | Death | T | Death1 | Death0 | 7.626 | <0.001 |
| Hypertension | ICU | T | ICU1 | ICU0 | 6.921 | <0.001 |
| | MV | F | MV1 | MV0 | 5.427 | <0.001 |
| | Death | F | Death1 | Death0 | 6.370 | <0.001 |

Table S9. Results of time-to-event prediction with Cox regression models

| | Cohort 2 (n = 682) | | | | | |
|---|---|---|---|---|---|---|
| | ICU | | MV | | Death | |
| Data | C Index | Integrated Brier Score | C Index | Integrated Brier Score | C Index | Integrated Brier Score |
| Radiom | 0.878 | 0.073 | 0.850 | 0.062 | 0.730 | 0.052 |
| RadioClinLab | 0.917 | 0.061 | 0.888 | 0.053 | 0.906 | 0.045 |
| ClinLab | 0.857 | 0.069 | 0.807 | 0.063 | 0.813 | 0.028 |
| | Cohort 3 (n = 652) | | | | | |
| | ICU | | MV | | Death | |
| Data | C Index | Integrated Brier Score | C Index | Integrated Brier Score | C Index | Integrated Brier Score |
| Radiom | 0.811 | 0.055 | 0.771 | 0.041 | 0.691 | 0.043 |
| RadioClinLab | 0.921 | 0.039 | 0.884 | 0.036 | 0.911 | 0.036 |
| ClinLab | 0.896 | 0.042 | 0.857 | 0.046 | 0.845 | 0.027 |

**Table S10. Bootstrapping results of Cox regression model for time-to-event prediction on Cohort 2 and Cohort 3**

| | Cohort 2 (n = 682) Bootstrapping Results | | | | | |
|---|---|---|---|---|---|---|
| | ICU | | MV | | Death | |
| Data | C Index (mean, 95% CI) | Integrated Brier Score (mean, 95% CI) | C Index (mean, 95% CI) | Integrated Brier Score (mean, 95% CI) | C Index (mean, 95% CI) | Integrated Brier Score (mean, 95% CI) |
| Radiom | 0.844 (0.795-0.893) | 0.095 (0.065-0.160) | 0.808 (0.731-0.863) | 0.076 (0.055-0.132) | 0.709 (0.618-0.792) | 0.059 (0.041-0.095) |
| RadioClinLab | 0.893 (0.792-0.935) | 0.077 (0.062-0.114) | 0.877 (0.824-0.930) | 0.066 (0.047-0.129) | 0.869 (0.775-0.905) | 0.046 (0.035-0.065) |
| ClinLab | 0.822 (0.761-0.876) | 0.075 (0.064-0.093) | 0.773 (0.679-0.880) | 0.071 (0.061-0.091) | 0.767 (0.624-0.911) | 0.042 (0.030-0.076) |

| | Cohort 3 (n = 652) Bootstrapping Results | | | | | |
|---|---|---|---|---|---|---|
| | ICU | | MV | | Death | |
| Data | C Index (mean, 95% CI) | Integrated Brier Score (mean, 95% CI) | C Index (mean, 95% CI) | Integrated Brier Score (mean, 95% CI) | C Index (mean, 95% CI) | Integrated Brier Score (mean, 95% CI) |
| Radiom | 0.783 (0.734-0.845) | 0.072 (0.039-0.133) | 0.728 (0.639-0.808) | 0.058 (0.034-0.131) | 0.686 (0.591-0.771) | 0.046 (0.038-0.064) |
| RadioClinLab | 0.885 (0.840-0.931) | 0.061 (0.037-0.134) | 0.877 (0.822-0.939) | 0.047 (0.030-0.152) | 0.870 (0.769-0.929) | 0.038 (0.032-0.065) |
| ClinLab | 0.877 (0.813-0.935) | 0.046 (0.033-0.070) | 0.831 (0.751-0.894) | 0.048 (0.036-0.059) | 0.790 (0.643-0.908) | 0.033 (0.026-0.052) |
