## Supplement Appendix 1-5 for "CT-based Rapid Triage of COVID-19 Patients: Risk Prediction and Progression Estimation of ICU Admission, Mechanical Ventilation, and Death of Hospitalized Patients"

**Appendix 1 Data collection**

Our multi-modal data included clinical data (clinical records and laboratory results), CT images, and radiologists’ semantic features. And the follow-up outcomes with time intervals of each patient.

(1) Clinical records (abbreviated as Clin): (a) demographics: age and gender; (b) comorbidities: coronary heart disease, diabetes, hypertension, chronic obstructive lung disease (COPD), chronic liver disease, chronic kidney disease, and carcinoma; and (c) clinical symptoms: fever, cough, myalgia, fatigue, headache, nausea or vomiting, diarrhea, abdominal pain, and dyspnea.

(2) Laboratory results (abbreviated as Lab): (a) blood routine: white blood cell (WBC) count (× 10⁹/L), neutrophil count (× 10⁹/L), lymphocyte count (× 10⁹/L), platelet count (× 10⁹/L), and hemoglobin (g/L); (b) coagulation function: prothrombin time (PT) (s), activated partial thromboplastin time (aPTT) (s), and D-dimer (mg/L); (c) blood biochemistry: albumin (g/L), alanine aminotransferase (ALT) (U/L), aspartate Aminotransferase (AST) (U/L), total bilirubin (mmol/L), serum potassium (mmol/L), sodium (mmol/L), creatinine (μmol/L), creatine kinase (CK) (U/L), lactate dehydrogenase (LDH) (U/L), α-Hydroxybutyrate dehydrogenase (HBDH) (U/L); (d) infection-related biomarkers: C-reactive protein (CRP) (mg/L).

For the laboratory data, we removed unambiguous extreme values with unclear units and test results with a missing rate of more than 50%, then performed median imputation on the remaining missing values (Table S2).

(3) CT-based radiomics features (abbreviated as Radiom): a total of 9943 quantitative radiomics features were extracted from CT images for each patient. First, 1657 radiomics features were extracted from each lesion. Next, for a given patient, for each feature, we summarized the distribution of the feature’s values across all the lesions for the patient by summary statistics (the mean, median, standard deviation, skewness, quartile 1, quartile 3), and obtained 1657*6 features per patient. We also add lesion count as a feature for each patient.

Chest CT scans were performed using ≥ 16 slice multidetector CT scanners (Aquilion ONE / Aquilion PRIME / BrightSpeed / BrightSpeed S / Brilliance 16 / Brilliance 64 / Discovery CT750 HD / eCT / Fluorospot Compact FD / HiSpeed Dual / iCT 256 / Ingenuity CT / Ingenuity Flex / LightSpeed VCT / LightSpeed 16 / NeuViz 16 Classic / Optima CT520 Series / Optima CT540 / Optima CT680 Series / ScintCare CT 16E / Sensation 64 / SOMATOM Definition AS+ / SOMATOM Definition Flash / uCT 510) without use of iodinated contrast agents. To minimize motion artifacts, patients were asked to hold their breath, then axial CT images were acquired during end-inspiration. The CT scan protocols were as follows: tube voltage, 100-120 kVp; effective tube current, 110-250 mAs; detector collimation, 16-320 x 0.625-2.5 mm; slice thickness, 0.625-2.5 mm; pitch, 0.8-1.375. Based on the raw data, the CT images were reconstructed by the iterative reconstruction technique reference needed.

The pneumonia was detected and segmented by a deep-learning AI system (Figure S2, Beijing Deepwise & League of PhD Technology Co.Ltd), which was built on top of deep convolutional neural networks and proved the performance by previous studies of COVID-19.^1, 2, 3^ Three major modules were designed to ensure the final accuracy of this system, i.e., pneumonia lesion detection, pneumonia lesion segmentation and lung lobe segmentation. First, an MVP-Net^4^ inspired method was used to detect bounding boxes of pneumonia findings. Channel-wise attention mechanism and multiple inputs (different windows centers and windows widths) were applied to explore the spatial context of the pneumonia in order to promise the detect sensitivity, and multiple symptom classifiers were trained to discriminate consolidation, ground-glass opacity, nodules and so forth. Pneumonia lesion (Figure S2), i.e., voxels that contained pneumonia, were extracted by a 3D U-Net.^5^ Finally, an anatomical prior embedded network was trained to partition the lung into five pulmonary lobes.^6^ Two practicing radiologists (C.S.Z and Q.M.X) randomly checked 100 cases and confirmed the effectiveness of the system.

Pyradiomics (v3.0) running in the Linux platform was adopted to extract radiomic features. Specifically, the radiomics features consisted of lesion count and 6 types of summary statistics of 1,657 original radiomics features extracted from each lesion. The original radiomics features comprised 19 First Order Statistics, 16 Shape-based (3D), 22 Gray Level Cooccurrence Matrix, 16 Gray Level Run Length Matrix, 16 Gray Level Size Zone Matrix, 14 Gray Level Dependence Matrix, 5 Neighboring Gray Tone Difference Matrix features, while the image type consisted of Original, Wavelet, LoG, Square, Logarithm, Gradient and LocalBinaryPattern3D. Then we extracted statistics (mean, standard deviation, median, skewness, quartile 1, quartile 3 of the lesion features, and the lesion count) to associate different lesions with related patients. Finally, a total of 9943 quantitative radiomics features were extracted from CT images for each patient.

(4) Radiologists’ semantic data (abbreviated as R-score): qualitative CT image features were evaluated by 10 experienced radiologists (5 to 18 years of experience). Our semantic features included (a) lesion distribution: subpleural or diffuse; (b) lesion morphology: round or other; (c) main signs: the presence of pure ground-glass opacity (GGO), pure consolidation, GGO with consolidation, interstitial lung disease (ILD), and crazy-paving pattern, (d) other abnormality: pleural effusion; and (e) the total number of lesions and lesion count in each lobe per patient.

All Digital Imaging and Communications in Medicine (DICOM) images were reviewed by four radiologists (Z.Y.S, L.Q, F. X., J. Z.) with 18, 6, 5, 5 years of experiences in thoracic radiology in core lab in Jinling Hospital, Medical School of Nanjing University. They independently assessed the axial CT images and/or multiplanar reconstructed images without access to the clinical or laboratory results of patients. The following features of chest thin-slice CT image were recorded: (a) lesion distribution: subpleural, diffuse or others; (b) lesion morphology: round, irregular or mixed; (c) main signs: the presence of pure ground-glass opacity (GGO), pure consolidation, GGO with consolidation, interstitial lung disease (ILD), crazy-paving pattern, (d) other abnormalities: fibrosis, pleural effusion, pulmonary emphysema, pulmonary edema. The total number of lesions and the number of lesions in each lobe per patient were also recorded.

GGO is defined as a fuzzy increase in lung attenuation without obscuring the underlying blood vessels.^7^ Consolidation is defined as increased attenuation of the lung parenchyma, blurring the edges of blood vessels and airway walls.^8^ ILD is defined as some sparing of individual lobules, forming a geographic-like appearance under the background of GGO, or the distortion of the lung structure and reticular opacities.^8^ Crazy-paving pattern is defined as thickened interlobular septa and intralobular lines on the background of ground glass turbidity.^8^ Subpleural distribution is defined as the lesion involving the peripheral 1/3 of the lung, while diffuse distribution is defined as continuous involvement without respect to lung segments.^7, 9^

Lungs are classified into five lung lobes. The left lung is divided into upper and lower lobes by oblique fissure, while the right lung is divided into upper, middle, and lower lobes by horizontal fissure and oblique fissure. We counted and recorded the number of lesions in each lobe: 0: no lesion; 1-5: with lesion and the number referred to the lesion number, regardless of the degree and range of the lesions (the highest number is 5). Consensus agreement was achieved through repeated examinations among radiologists.

(5) Time-to-event data: we gathered the following follow-up data. The time intervals between the date of admission and (a) the date of development of adverse outcomes (requiring ICU, MV, and death), or (b) the date of discharge (defined as a patient who had no fever for at least 3 days, a significant improvement on chest CT in both lungs, clinical relief of respiratory symptoms, and repeated negative RT-PCR results at ≥ 24 hours interval10) were recorded.

**Appendix 2 Feature processing methods**

Several feature engineering methods were applied: 1) SMOTEENN (synthetic minority over-sampling technique and edited nearest neighbors):^11^ The method performs over-sampling using SMOTE and cleaning using ENN to deal with imbalanced classes. In this study, a 1:1 (positive cases: negative cases) balanced dataset and a 1:3 imbalanced dataset were created respectively; 2) SMOTEENN+PCA (principal component analysis):^12, 13, 14^ upon enlarging the dataset, PCA was applied to reduce the dimensionality of the features. It applies singular value decomposition (SVD) to find the orthogonal principal components and the low-dimension representation of data. In this study, the number of principal components was chosen to explain 0.998 or 0.954 variance; 3) SMOTENN + LASSO feature selection:^15^ LASSO feature selection was applied to extract the most important features used in logistic regression with L1 normalization. coefficients of L1 normalization (‘C’) were tuned; 4) SMOTEENN+GUS (generic univariate selection):^16^ Generic univariate selection selects the best features based on univariate statistical tests; 5) SMOTENN+FPR (false positive rate test).^16^ The feature engineering was done with the toolbox of scikit-learn 0.23.0.^16^ In our study, for the last two feature selection methods, F-test and mutual information were used as the scoring function. The feature selection was done with the scikit-learn 0.23.0.^16^ The modeling process was done with the raw data and preprocessed data with the methods mentioned above.

**Appendix 3 Feature visualization**

Feature visualization provides an intuitive manner to understand the distribution of features used in this study. Therefore, we first visualized the distribution of 37 clinical data (including 18 clinical features and 19 laboratory test results), 9,943 CT-based radiomics features, and 17 traditional semantic CT features for all patients, with the help of heatmaps and t-distributed Stochastic Neighbor Embedding (t-SNE) in terms of ICU, MV, and death (ComplexHeatmap version 2.2.0).^17, 18^ The patients were reasonably grouped based on the adverse outcomes and whether the event occurred within 48 hours.

We recognized the marked differences of radiomics data, clinical data, and R-score data on Cohort 1 and Cohort 2 between negative outcome patients and positive outcome patients (Figure 2, Figure S3). Almost all CT image features showed good discrimination between negative and severe outcome patients and had more obvious distinctions compared to clinical data. Among clinical data, lab results and demographics had good discriminating power. Part of radiologists’ score features had good discriminating power while clinical features have comparatively weak discriminating power. Regarding the distinctions between ICU admission, mechanical ventilation, and death, CT image features showed better discriminating power than clinical data. In CT image features, from ICU to MV to death, trends of value increasing or decreasing can be observed while in clinical data, this kind of trend is not visible. All in all, we expect better learning results with features of better discriminating power, and the feature and model analysis results showed this expectation.

**Appendix 4 Machine-learning classifiers**

We used the following approaches to systematically explore the performance of multiple machine-learning classifiers to predict outcomes (ICU, MV, and death): 1) Logistic Regression (LR);^19^ 2) Random Forest (RF);^20^ 3) Support Vector Machine (SVM);^21^ 4) Multilayer Perceptron (MLP);^22^ 5) LightGBM.^23^ The hyperparameters tuned for each of the algorithms included: 1) LR: the coefficient of L2 normalization (‘C’); 2) RF: the number of estimators (‘n_estimators’), maximum depth (‘max_depth’); 3) SVM: the coefficient of soft margin relaxation (‘C’) with the radial basis function kernel; 4) MLP: the number of hidden units in a two-layer fully connected neural network; 5) LightGBM: learning rate, the number of estimators (‘n_estimators’), the number of leaves(‘num_leaves’).

**Appendix 5 Key imaging features and clinical prognostic indicators**

Among the prognostic indicators chosen based on bootstrapping experiments with top-ranking feature importance used in the optimal models for the three prediction tasks (RadioClinLab), clinical data and radiomics features showed a complementary role with no significant correlations (Figure 3, Figure S5-6). In clinical data, elder age, dyspnea, higher lactate dehydrogenase (LDH) and inflammatory factors (white blood cell (WBC), neutrophil) signaled severe outcomes. Particularly, hypertension and other inflammatory factors (lower lymphocyte, higher C-reactive protein (CRP) and neutrophil)) were valuable for predicting ICU admission, also potassium which related to acid-base and electrolyte disorders, the indicator of myocardial infarction (higher α-Hydroxybutyrate dehydrogenase (HBDH)) and several inflammatory factors (lower lymphocyte, higher CRP) were predictive for MV, while higher D-dimer provided great diagnostic value for death. Furthermore, GLSZM-based, GLCM-based, and first-order radiomics features are important features for the prediction of outcomes. In addition, the top-ranking traditional CT features that contribute most to the R-score model suggested that diffuse pulmonary parenchymal ground-glass and consolidative pulmonary opacities in the left upper lobe and pleural effusion increased the adverse outcomes (ICU, MV, death) in COVID-19 patients. Notably, crazy-paving on the initial CT chest was a risk factor of death. (Table S6, Figure S7)

The statistical difference between negative and positive cases on the majority of the ten most important features (10/10 for ICU prediction, 10/10 for MV prediction, 9/10 for death prediction) in three outcome prediction tasks on Cohort 1 was confirmed (Table S7). On Cohort 2, the feature values of the negative and positive cases on the majority of the ten most important features (10/10 for ICU prediction, 9/10 for MV prediction, 9/10 for death prediction) found by classifiers also showed statistically significant difference (Table S8). The reason that there were no significant differences in some of the lab results might be that the imputation of the missing values was based on median values on Cohort 1 and the standardization was done based on Cohort 1 (training data). Therefore, due to the distributional differences, the statistical significance was not evident in all the features.

**Appendix Table and Figure captions**

**Table captions**

Table S1. Geographical distribution of COVID-19 patients in this study

Table S2. Characteristics of patients in stable/adverse (ICU) groups, non-MV/MV groups, and discharge/death groups

Table S3. Statistical significance test of top ten important feature values of positive cases between Cohort 1 and Cohort 2

Table S4. Performance and algorithms of the optimal models of each data type for the prediction of ICU, MV, and death on the three cohorts

Table S5. Statistical significance of the bootstrapping results of different data modalities in Cohort 2 and Cohort 3

Table S6. The feature weight of traditional CT-based features on three outcome prediction tasks

Table S7. The statistical significance of the difference between negative and positive cases on all of the ten most important features in three outcome prediction tasks on Cohort 1

Table S8. The statistical significance of the difference between negative and positive cases on all of the ten most important features in three outcome prediction tasks on Cohort 2

Table S9. Results of time-to-event prediction with Cox regression models

Table S10. Bootstrapping results of Cox regression model for time-to-event prediction on Cohort 2 and Cohort 2 Subset

**Figure captions**

**Figure S1. Geographical distribution of COVID-19 patients in this study. (a)** COVID-19 patients (n = 2362) were analyzed from 26 hospitals in 14 provinces in China (including Hubei, Anhui, Chongqing, Hunan, Henan, Hainan, Jiangxi, Zhejiang, Jiangsu, Haerbin, Shandong, Liaoning, Xinjiang, and Guizhou). **(b)** patients (n = 1721) enrolled in each city in Hubei Province (including Wuhan, Huangshi, Huanggang, Jingzhou, Xiaogan, Yichang, Xiangyang).

**Figure S2. Flowchart and example segmentations of the AI system. (a) Flowchart of the AI system. (a-f) Examples of lesion segmentation by the AI system.** Left (a, c, e): original images; Right (b, d, f): pulmonary lobes (colored lines) and opacities segmentation (blue area).

**Figure S3. t-SNE visualization of cases in Cohort 1 with five different combinations of data types.** Each point denotes a patient, where yellow represents patients discharge without any adverse outcome and blue represents patients with events for ICU, MV, or death. The distance between each point is positively related to the similarity of that lesion of patients. Note: with only the raw data, the positive cases and negative cases are not visually well separated. This calls for necessary feature engineering steps and classifiers to select features important to classification.

**Figure S4. The box plots of bootstrapping model performances in terms of AUPRC and AUPRC on three outcome prediction tasks on Cohort 1 (a-f) and Cohort 2 (g-l).**

**Figure S5. The paired plot of the ten most important features in (a) ICU, (b) MV, (c)death prediction.** Most important features are respectively: (the ranks align with the plots) (a) ICU: 1.Dyspnea, 2.Age, 3.LDH, 4.wavelet-LHH_glszm, 5.WBC, 6.original_glszm, 7.lymphocyte, 8.CRP, 9.hypertension, 10.neutrophil; (b) MV: 1.Dyspnea, 2.Neutrophil, 3.Age, 4.LDH, 5.CRP, 6.HBDH, 7.Lymphocyte, 8.Potassium, 9.WBC, 10.wavelet-LHH_glszm; (c) Death: 1.Dyspnea, 2.LDH, 3.Age, 4.WBC, 5.wavelet-HLH_glcm, 6.original_firstorder, 7.wavelet-LHL_firstorder, 8.Neutrophil, 9. wavelet-HLL_glszm, 10. D-dimer. Plots at the Diagonal positions are histograms of the most important features while plots at non-diagonal positions are scatter plots of a pair of most important features. Each point represents a patient in Cohort 2. No significant correlations are found between radiomics features and clinical features.

**Figure S6. The feature values of Cohort 1 patients with and without adverse outcomes of the continuous features among all the ten most important features found on three prediction tasks.** The feature values of positive and negative cases of the fifteen continuous variables found among the top ten important features on three tasks (Cohort 1 samples) were shown. The statistical significance was tested and shown (Table S7) on the respective ten most important features (both continuous and categorical variables) in ICU prediction, MV prediction and death prediction.

**Figure S7. Examples of semantic features on the initial CT scan of COVID-19 patients with severe outcomes.** (a) Axial CT image obtained without enhancement in a 60-year-old man shows consolidative opacity with a rounded morphology (white arrow) in right lower lobe and shows consolidative opacity with peripheral distribution in left lower lobe (black arrow). (b) Axial CT image obtained in a 60-year-old man shows consolidative opacities with peripheral distribution in the right lower lobe (arrows). (c) Axial CT image without intravenous contrast material in a 47-year-old woman shows bilateral ground-glass and consolidative opacities with a striking peripheral distribution (arrows). (d) Axial CT image obtained in a 39-year-old man shows ground-glass opacities and consolidative opacities with diffuse distribution(arrows).

**References**

1 Ni Q, Sun ZY, Qi L, et al. A deep learning approach to characterize 2019 coronavirus disease (COVID-19) pneumonia in chest CT images. Eur Radiol 2020;1-11.

2 Yu Q, Wang Y, Huang S, et al. Multicenter cohort study demonstrates more consolidation in upper lungs on initial CT increases the risk of adverse clinical outcome in COVID-19 patients. Theranostics 2020;10:5641-5648.

3 Wang YC, Luo HY, Liu SQ, et al. Dynamic evolution of COVID-19 on chest computed tomography: experience from Jiangsu Province of China. Eur Radiol 2020; https://doi.org/10.1007/s00330-020-06976-6.

4 Li ZH, Zhang S, Zhang J, Huang KQ, Wang YZ, Yu YZ. MVP-Net: Multi-view FPN with Position-aware Attention for Deep Universal Lesion Detection. International Conference on Medical Image Computing and Computer-Assisted Intervention 2019; arXiv:1909.04247.

5 Huang C, Wang Y, Li X, et al. Clinical features of patients infected with 2019 novel coronavirus in Wuhan, China. Lancet 2020; 395, 497-506.

6 Guan WJ, Ni ZY, Hu Y, et al. Clinical Characteristics of Coronavirus Disease 2019 in China. New England Journal of Medicine 2020; 10.1056/NEJMoa2002032.

7 Song F, Shi N, Shan F, et al. Emerging coronavirus 2019-nCoV pneumonia. Radiology. 2020; 295: 210-7.

8 Hansell DM, Bankier AA, MacMahon H, McLoud TC, Muller NL, Remy J. Fleischner Society: glossary of terms for thoracic imaging. Radiology 2008; 246:697-722.

9 Ooi GC, Khong PL, Müller NL, et al. Severe acute respiratory syndrome: temporal lung changes at thin-section CT in 30 patients. Radiology. 2004; 230:836-44.

10 Zhou F, Yu T, Du R, et al. Clinical course and risk factors for mortality of adult inpatients with COVID-19 in Wuhan, China: a retrospective cohort study. The Lancet. 2020; 395(10229): 1054-1062.

11 Batista GE, Ana LCB, Monard MC. Balancing Training Data for Automated Annotation of Keywords: a Case Study. WOB 2003.

12 Pearson, Karl. LIII. On lines and planes of closest fit to systems of points in space. The London, Edinburgh, and Dublin Philosophical Magazine and Journal of Science 2010; 2:11, 559-572.

13 Jirsa VK, Friedrich R, Haken H, et al. A theoretical model of phase transitions in the human brain. Biological cybernetics 1994; 71.1: 27-35.

14 Zhan XH, Guan XQ, Wu RM, Wang Z, Wang Y, Li G. Discrimination between Alternative Herbal Medicines from Different Categories with the Electronic Nose. Sensors(Basel) 2018; 18(9): 2936.

15 Tibshirani, Robert. Regression shrinkage and selection via the lasso. Journal of the Royal Statistical Society: Series B (Methodological) 1996; 58.1: 267-288.

16 Fabian P, Varoquaux G, Gramfort A, Michel V, Thirion B. Scikit-learn: Machine learning in Python. Journal of Machine Learning Research 2011; 12: 2825-2830.

17 Gu Z, Eils R, Schlesner M. Complex heatmaps reveal patterns and correlation in multidimensional genomic data. Bioinformatics 2016; 15;32(18):2847-9

18 Maaten L, Hinton G. Visualizing data using t-SNE. Journal of machine learning research 2008; 9:2579-2605.

19 Cramer JS. The Origins of Logistic Regression. Tinbergen Institute, Tinbergen Institute Discussion Papers 2002; 10.2139/ssrn.360300.

20 Ho TK. Random decision forests. IEEE 1995; 1:278-282.

21 Ben-Hur A, Horn D, Siegelmann HT, Vapnik V. Support vector clustering. J. Mach. Learn. Res. 2 2001; 125–137.

22 Collobert R and Bengio S. Links between Perceptrons, MLPs and SVMs. ICML 2004; 04.

23 Ke G, Meng Q, Finley T, et al. Lightgbm: A highly efficient gradient boosting decision tree. Advances in neural information processing systems 2017; <https://papers.nips.cc/paper/6907-lightgbm-a-highly-efficient-gradient-boosting-decision-tree.pdf.>
